# Impact of the COVID-19 Pandemic on Personal Networks and Neurological Outcomes of People with Multiple Sclerosis: A Case-Control Cross-sectional and Longitudinal Analysis

**DOI:** 10.1101/2022.08.17.22278896

**Authors:** Claire S. Riley, Shruthi Venkatesh, Amar Dhand, Nandini Doshi, Katelyn Kavak, Elle E. Levit, Christopher Perrone, Bianca Weinstock-Guttman, Erin E. Longbrake, MSReCOV investigators, Philip L. De Jager, Zongqi Xia

**Affiliations:** Multiple Sclerosis Center and the Center for Translational & Computational NeuroImmunology, Department of Neurology, Columbia University Irving Medical Center, New York, NY; Department of Neurology, University of Pittsburgh School of Medicine, Pittsburgh, PA; Department of Neurology, Division of Hospital Medicine, Division of Stroke and Cerebrovascular Diseases, Brigham and Women’s Hospital, Boston, MA; Harvard Medical School, Boston, MA; Jacobs MS Center for Treatment, Jacobs Medical School and Biomedical Sciences, SUNY at Buffalo; Department of Neurology, Yale School of Medicine, Yale University, New Haven, CT; Department of Neurology, University of Pennsylvania, Philadelphia, PA

## Abstract

**Background:** The COVID-19 pandemic has negatively impacted the social fabric of people with multiple sclerosis (pwMS).

**Objective:** To evaluate the associations between personal social network environment and neurological function in pwMS and controls during the COVID-19 pandemic and compare with the pre-pandemic baseline.

**Methods:** We first analyzed data collected from 8 cohorts of pwMS and control participants during the COVID-19 pandemic (March-December 2020). We then leveraged data collected between 2017-2019 in 3 of the 8 cohorts for longitudinal comparison. Participants completed a questionnaire that quantified the structure and composition of their personal social network, including the health behaviors of network members. We assessed neurological disability using three interrelated patient-reported outcomes: Patient Determined Disease Steps (PDDS), Multiple Sclerosis Rating Scale – Revised (MSRS-R), and Patient Reported Outcomes Measurement Information System (PROMIS)-Physical Function. We identified the network features associated with neurologic disability using paired t-tests and covariate-adjusted regressions.

**Results:** In the cross-sectional analysis of the pandemic data from 1130 pwMS and 1250 control participants, higher percent of network members with a perceived negative health influence was associated with greater neurological symptom burden in pwMS (MSRS-R: Beta[95% CI]=2.181[1.082, 3.279], p<.001) and worse physical function in controls (PROMIS-Physical Function: Beta[95% CI]=-5.707[-7.405, -4.010], p<.001). In the longitudinal analysis of 230 pwMS and 136 control participants, the networks of both pwMS and controls experienced an increase in constraint (pwMS p=.006, control p=.001) as well as a decrease in network size (pwMS p=.003, control p<.001), effective size (pwMS p=.007, control p=.013), maximum degree (pwMS p=.01, control p<.001), and percent contacted weekly or less (pwMS p<.001, control p<.001), suggesting overall network contraction during the COVID-19 pandemic. There was also an increase in percentage of kin (p=.003) in the networks of pwMS but not controls during the COVID-19 pandemic when compared to the pre-pandemic baseline. These changes in personal social network due to the pandemic were not associated with worsening neurological disability during the pandemic.

**Conclusions:** Our findings suggest that perceived negative health influences in personal social networks are associated with worse disability in all participants during the COVID-19 pandemic. Despite the perturbation in social environment and connections during the pandemic, the stability in neurological function among pwMS suggests potential resilience.

## INTRODUCTION

Multiple sclerosis (MS) is a chronic autoimmune disease affecting the central nervous system and causing neurodegeneration and neurological disability.^1, 2^ While a multitude of factors potentially influence MS susceptibility, such as genetic variants, cigarette smoking, Epstein-Barr virus infection, adolescent obesity, and Vitamin D deficiency, factors that influence disease progression are less well defined.^3^ Studies of personal social networks raise intriguing possibilities that social environments impact health outcomes and neurological function.^4^

Personal social network features are associated with the quality of life of people living with neurological diseases, including stroke, traumatic brain injury, and MS.^5–7^ Our prior research implicates the personal social network as a potentially modifiable environmental contributor to neurological disability in people with MS (pwMS).^8, 9^ Adverse personal social networks are associated with social isolation and loneliness, which may have direct biological effects by altering inflammation, recovery from injury, and resilience against neurodegeneration.^10–13^ Personal social network structures (*e.g.,* larger network size, more diffuse connections among network members) are associated with language function and regional brain volume in pwMS.^14^ Examining personal social network structure (*e.g.,* network size, density) and composition (*e.g.,* demographics, health behaviors of the network members) in relation to disability accumulation in MS could potentially inform novel interventions. Further, pwMS are often more vulnerable to isolation and loneliness than the general population and have experienced more pronounced negative impacts during the coronavirus disease 2019 (COVID-19) pandemic partially due to their greater need for social distancing.^15–17^

In this study, we assessed the personal social networks of pwMS and control participants during the COVID-19 pandemic and identified network features associated with worse neurological outcomes. Additionally, we examined changes in personal social networks due to the COVID-19 pandemic in a subset of previously characterized individuals.^8, 9^

## METHODS

### Study Cohort

For cross-sectional analyses, we included pwMS and healthy control participants from clinic-based cohorts at University of Pittsburgh Medical Center (UPMC, 2 cohorts)^18, 19^, Columbia University Irving Medical Center (CUIMC, 2 cohorts), Yale University, University of Pennsylvania, and University of Buffalo recruited between March and December 2020 through the Multiple Sclerosis Resilience to COVID-19 (MSReCOV) Collaborative.^15–17^ Additionally, we included participants from a national cohort of first degree relatives of MS patients (Genes and Environment in Multiple Sclerosis [GEMS] Study), including both pwMS and controls.^20, 21^ The inclusion criteria were adults aged 18 or older either with or without a neurologist-confirmed diagnosis of MS. We deployed a modified personal network questionnaire (PERSNET, see Supplementary Material) to assess demographic, clinical and personal social network features through a secure online platform (Research Electronic Data Capture).^22^ For longitudinal analyses, we leveraged PERSNET data collected prior to the COVID-19 pandemic (2017-2019) from UPMC, CUIMC, and the GEMS cohorts (**Figure 1A**).^9^

**Figure 1.**
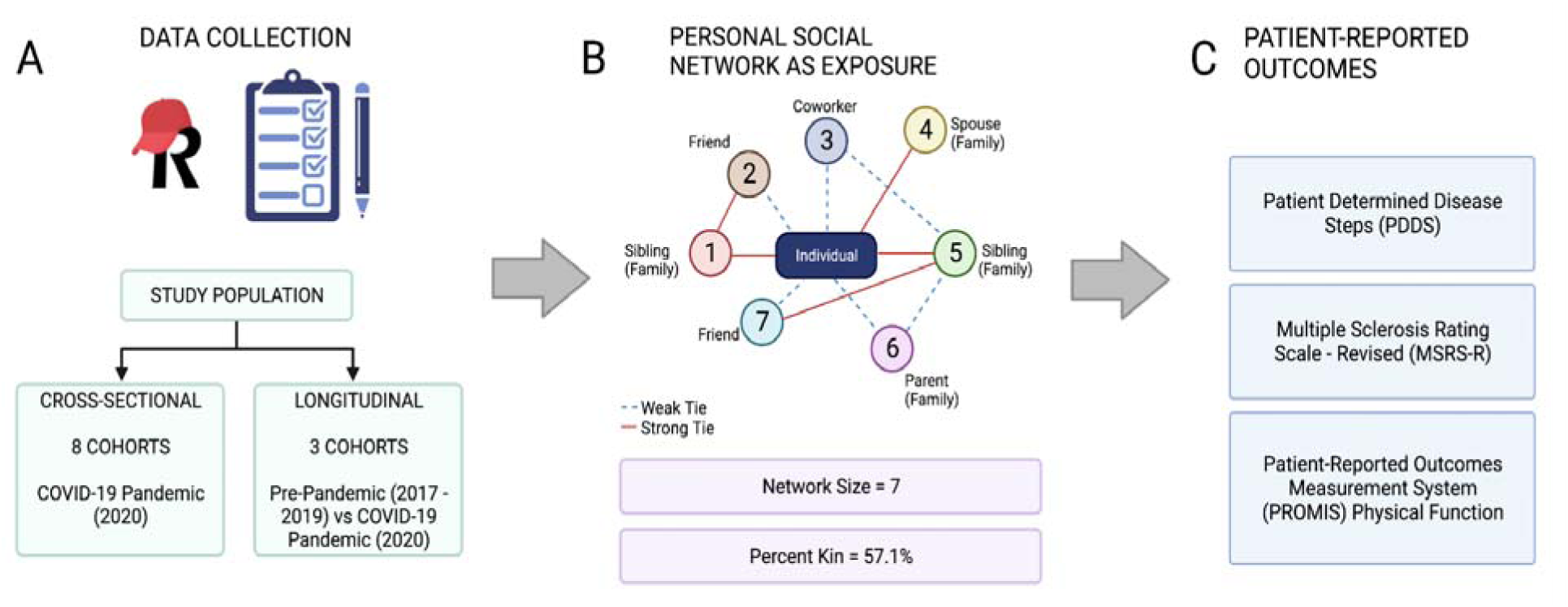
Overview of study design. A: schematic illustration of study populations, survey data collection using <u>R</u>esearch <u>E</u>lectronic <u>D</u>ata <u>Cap</u>ture (REDCap); B: personal social network features as exposure, as illustrated by a representative network of a hypothetical participant; C: patient-reported outcomes of neurological disability or physical function.

### Ethics Approval

The institutional review board of each enrolling site approved the study. All participants provided informed consent.

### Personal Network Metrics

We deployed an updated version of the PERSNET, adapted from the foundational General Social Survey.^5–9, 23^ PERSNET assesses two categories of features characterizing the interconnectedness of the relationships maintained by each person.^24, 25^

Network structure includes 6 quantitative features: size (number of individuals in the network, excluding the index person), density (sum of ties, excluding the index person’ ties, divided by all possible ties), constraint (a more granular density that assesses the extent to which the index person’s connections are to individuals who are connected to one another), effective size (number of non-redundant network members), maximum and mean degrees (highest and average number of ties by a network member).

Network composition quantifies demographic characteristics and health behaviors of network members. Network demographics include percent kin (percent of network members who are family), standard deviation (SD) of age (age range of network members), diversity of sex index (proportion of sexes from 0 to 1, where 0 indicates single sex and 1 indicates equal ratio of men and women), diversity of race (similar proportion of represented races where 0 indicates a single race). Network health behaviors include the percent of network members who smoke, consume alcohol, exhibit poor dietary habits, lead sedentary lifestyles, and exert perceived negative health influence. Other network composition features include the frequency, duration, and the living distance of social contact, which quantify the depth of the relationships. Compositional features account for network size.

### Neurological Outcomes

To assess neurological function, we used three interrelated patient-reported outcomes (PROs). Patient Determined Disease Steps (PDDS) score indicates the extent of the gait impairment and highly correlates with the clinician-determined Expanded Disability Status Scale (EDSS). PDDS scores range from 0 to 8 where 0 corresponds to normal while 8 indicates bed-bound status.^26^ Multiple Sclerosis Rating Scale-Revised (MSRS-R) assesses the global neurological symptom burden, including walking, function in the extremities, vision, speech, swallowing, cognition, sensation, bladder and bowel function.^27, 28^ Each symptom domain score ranges from 0 to 4 where 0 indicates no impairment and 4 indicates severe impairment. Higher cumulative MSRS-R scores (0-32) indicate greater neurological symptom burden and worse neurological function. Patient-Reported Outcomes Measurement Information System (PROMIS)-Physical Function (version 1.2) is a generalizable measure of physical function but also validated for pwMS.^29, 30^ PROMIS Physical Function is reported as normally distributed T-score on a 0-100 scale, where 50 represent the average for the US population and higher scores indicate better function. While PDDS and MSRS-R are specific for pwMS, PROMIS is applicable to both pwMS and controls. All participants completed PRO assessment when completing the PERSNET during the COVID-19 pandemic, while a subset also completed the PROs before the pandemic.

### Covariates

We considered the following confounding factors that could potentially influence neurological function: age, sex, race, ethnicity, disease duration, employment, education, occupation, income, marital status, and cohabitant status. Race was categorized as African or African American, American Indian or Native Hawaiian or Pacific Islander, Asian, White, Multi-racial or other. Ethnicity was categorized as Hispanic (or Latino) and Non-Hispanic. Due to the relatively small number of racial and ethnic minorities in our cohorts, race and ethnicity were dichotomized as Non-Hispanic White versus others (encompassing individuals of Hispanic and/or non-European descent) in subsequent analyses. Disease duration was defined as the time from the first self-reported neurological symptom onset to time of the most recent PRO assessment. Employment status was categorized as employed for wages, self-employed, out of work and looking for work, out of work but not currently looking for work, homemaker, student, military, retired, or unable to work. Education level was classified based on the highest level of education achieved: some high school or less, high school graduate, some college, associate degree, bachelor’s degree, or graduate degree. Occupation was categorized as business owner, executive/manager, professional, sales/clerical worker, service worker, or other. Household income level included the following brackets: 0 to $19,999, $20,000 to $34,999, $35,000 to $49,999, $50,000 to $64,999, $65,000 to $79,999, $80,000 to $94,999, $95,000 to $109,999, $110,000 to $124,999, or $125,000 or higher. Marital status was categorized as married or unmarried. Cohabitant status was categorized as living alone versus otherwise. Some covariates were missing because certain questions (*e.g.,* employment status, income) in PERSNET were made optional in order to reduce patient discomfort when completing the questionnaire.

To narrow the list of covariates, we assessed the correlation between these features and PROs in univariate analyses (**eFigure 1**). We selected features that met the following pre-defined criteria as covariates for downstream analyses: feature presence in over 70% of pwMS, a Pearson’s correlation coefficient of 0.1 or higher, and a nominal statistical significance (p<.05) in association with all three PROs. We identified age, disease duration, employment, and income as meeting the criteria and adjusted these covariates in regression models involving pwMS. For analyses involving control participants, we adjusted for age, employment, and income but not disease duration as it does not apply to controls. For longitudinal analyses, we further adjusted for the time elapsed between pre-pandemic and pandemic PERSNET assessment as well as study cohort as additional covariates for consistency with our previous analysis.^8, 9^

### Statistical Analysis

We performed two types of analyses: (a) cross-sectional analysis of pwMS and controls during the COVID-19 pandemic, (b) longitudinal analysis of pwMS and controls during the COVID-19 pandemic as compared to the pre-pandemic baseline.

For the cross-sectional analysis, we first compared the personal social networks of pwMS and control participants using paired T-tests. Next, we examined the association between structural and compositional network features and PROs in pwMS in covariate-adjusted regression models. Given that PROMIS measure is generalizable across health and disease, we further assessed the association between network features and PROMIS Physical Function in control participants for comparison. In a joint analysis of the pandemic data that include both pwMS and controls, we performed a moderation analysis to assess whether MS diagnosis influenced the association between network features and PROMIS Physical Function.^31–33^

For the longitudinal analysis, we examined the *within-subject* differences in network features in pwMS and control participants during the COVID-19 pandemic as compared to the most recent pre-pandemic baseline using paired T-tests. Next, we assessed the association of change in structural and compositional network features (*i.e.,* pandemic value minus pre-pandemic baseline) in relation to the most recent PROs (during the pandemic) in pwMS using covariate-adjusted regressions and an omnibus test.^9, 21^ For the omnibus test, we combined the p-values derived from the covariate-adjusted regressions for each PRO. Using Fisher’s combined probability test, we calculated the chi-squared statistic and compared the observed values to the expected empirical distribution. To additionally assess these relationships, we generated a quantile-quantile (Q-Q) plot of the observed versus expected p-values of the associations between the longitudinal changes (pandemic values minus pre-pandemic baseline) in each structural and compositional network feature and each PRO (during the pandemic). The 95% confidence intervals of the Q-Q plot were obtained from 10,000 permutations of the p-value distribution. We performed similar longitudinal analysis in control participants using PROMIS Physical Function for comparison. Raw p-values were adjusted by Bonferroni correction for multiple comparisons. All statistical analyses were performed using R version 3.6.0.^34^

### Code Availability

The code for this project is available at <https://github.com/shruthi1094/pnq>. De-identified data are available upon request to the corresponding author and with permission from each participating center.

## RESULTS

### Participant Characteristics

The cross-sectional analysis included 1130 pwMS aged (mean±SD) 50.7±12.1 and 1250 control participants aged 44.3±12.1 years (**Table 1**). Participants were predominantly women (81.9% pwMS, 76.8% controls) and Non-Hispanic White (87.1 pwMS, 86.7% controls). PwMS were less likely to be employed (48.1%) than controls (78.1%). The disability burden among pwMS was mild to moderate (mean±SD PDDS 1.85±2.12, MSRS-R 7.55±5.49, PROMIS Physical Function 46.4±10.82).

**Table 1.**
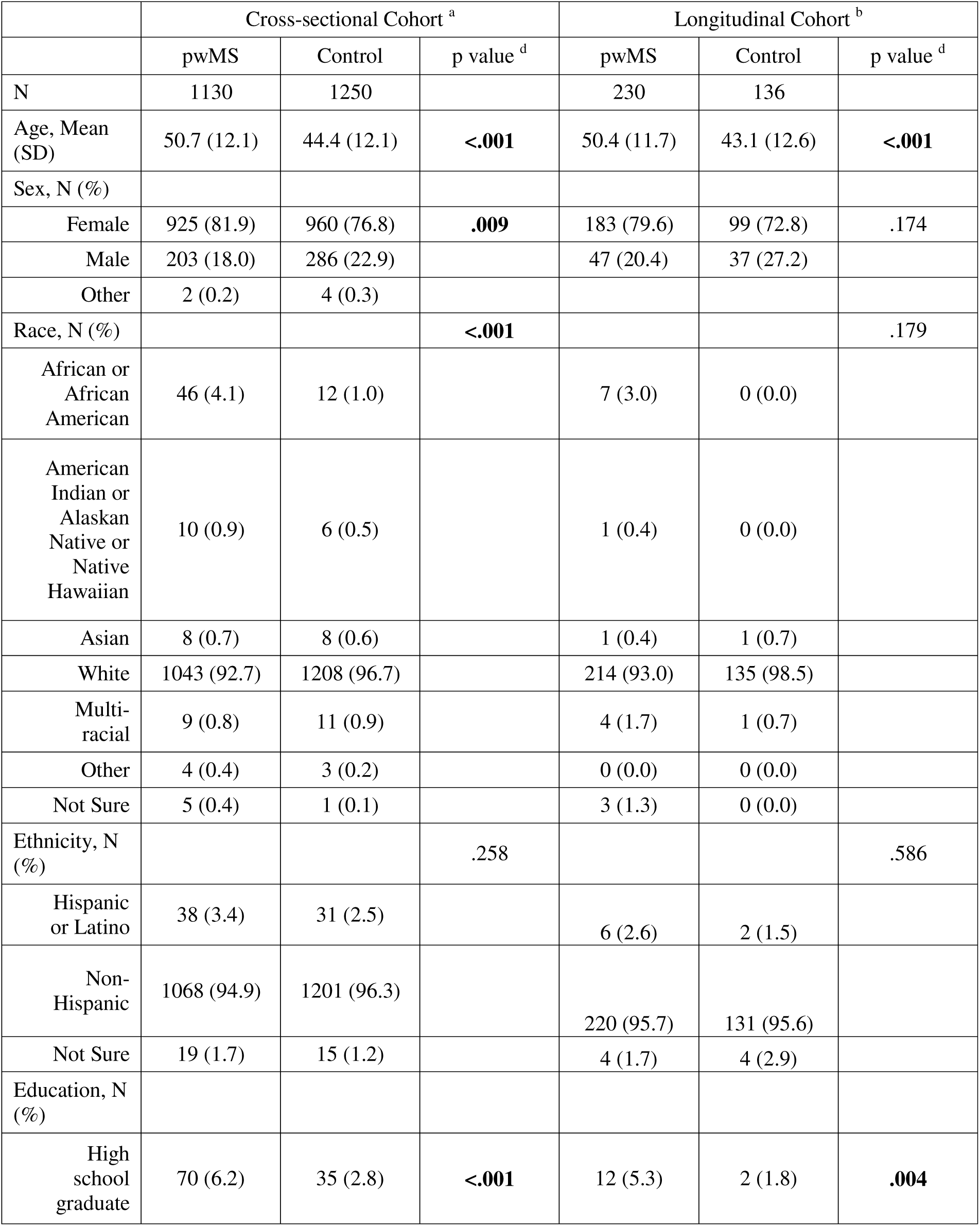
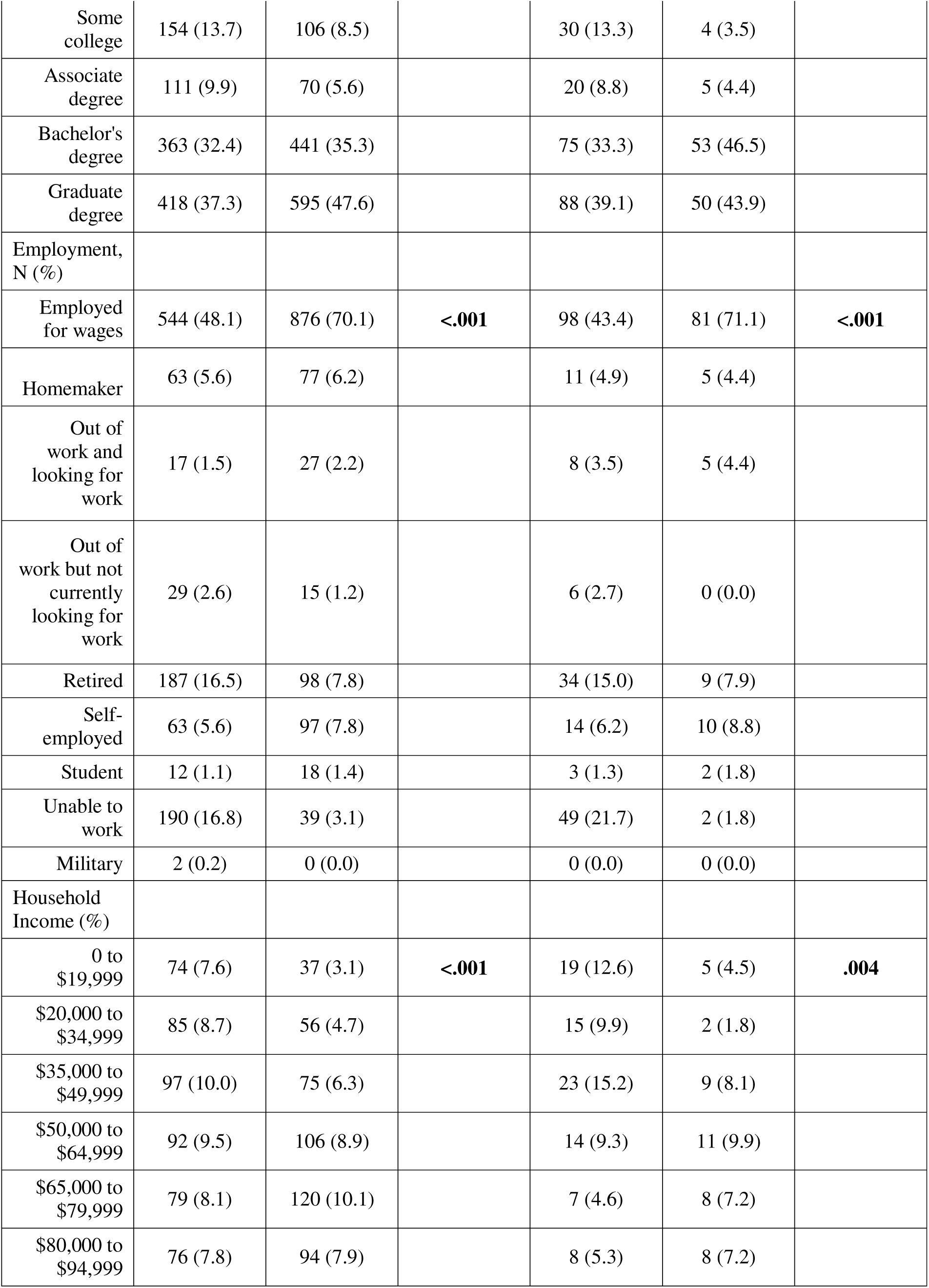
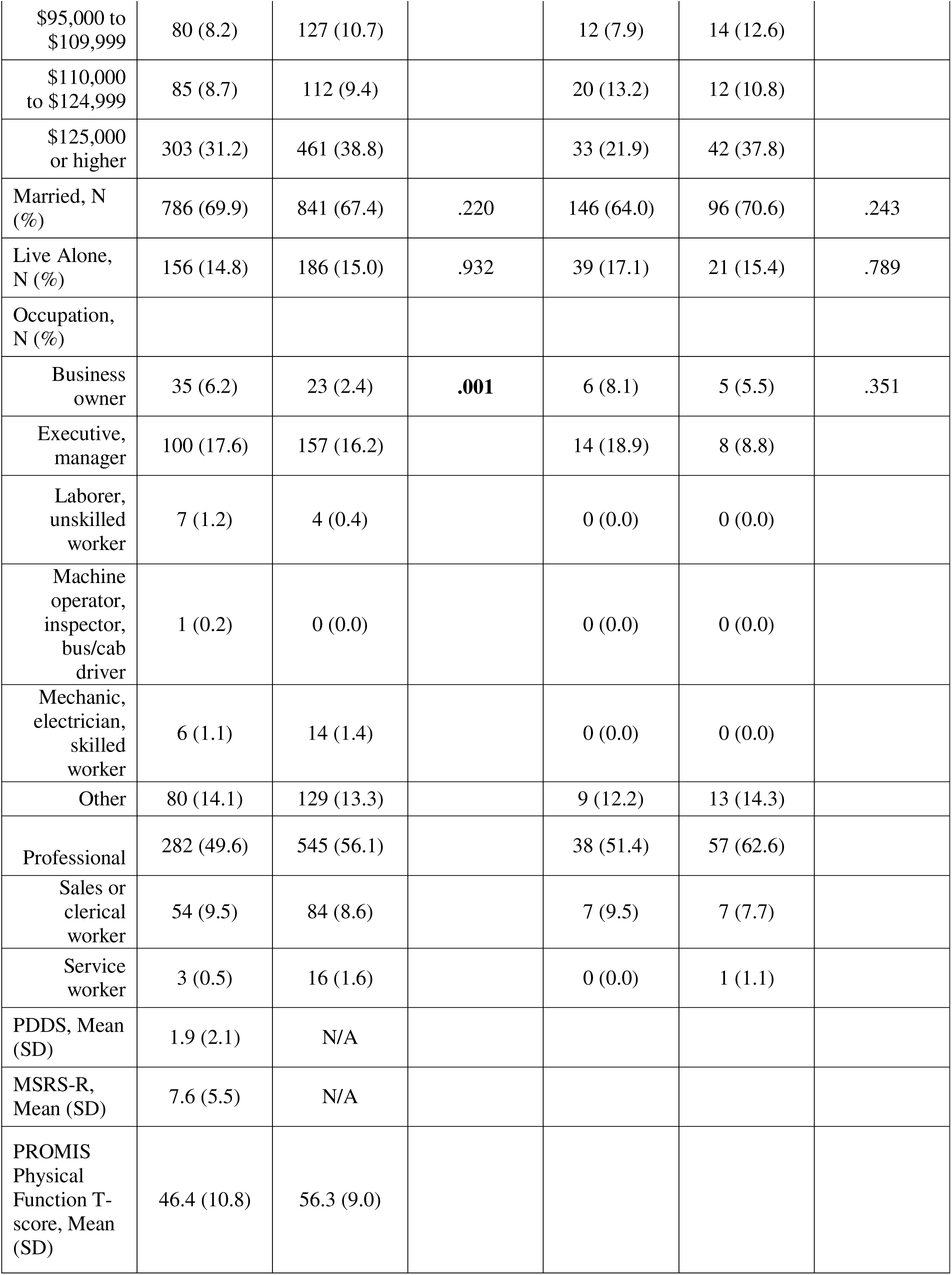
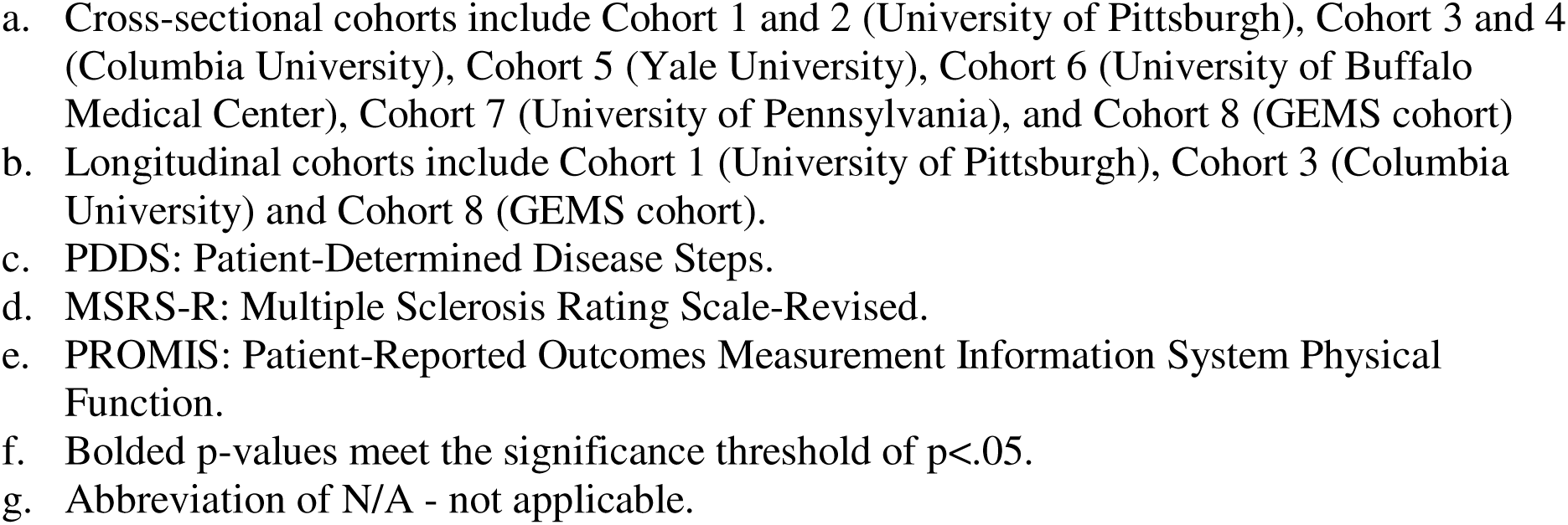
Participant characteristics of the cross-sectional and longitudinal cohorts.

The longitudinal analysis included 230 pwMS and 136 control participants. Likewise, control participants were younger (mean±SD pwMS: 50.4±11.7 years, controls: 43.1±12.6 years), had higher levels of education (81.2% pwMS and 94.8% controls completed college), were employed (43.4% pwMS, 71.1% controls) and had higher household income than pwMS.

### Cross-Sectional Analysis of the Pandemic Period

First, we compared the network features of pwMS and control participants during the COVID-19 pandemic (**Table 2**). In this unadjusted analysis, pwMS had higher density, higher constraint, smaller effective size, higher percent kin, lower percent of people known for less than 6 years, lower percent who live over 15 miles away, and lower percent who drink in their social networks when compared to control participants. PwMS and controls reported similarly high percent of their network members with a perceived negative health influence (36.1% pwMS, 36.3% control).

**Table 2.** Comparison of personal social network features in people with multiple sclerosis and control participants during the COVID-19 pandemic.

|  | pwMS<br>(N = 1130) | Control<br>(N = 1250) | p value <sup>a</sup> |
| --- | --- | --- | --- |
| *Network Structure* |  |  |  |
| Network Size, Mean (SD) | 6.79 (4.24) | 6.78 (3.90) | .946 |
| Density, Mean (SD) | 0.76 (0.25) | 0.72 (0.24) | **<.001** |
| Constraint, Mean (SD) | 56.88 (19.07) | 54.30 (17.03) | **.001** |
| Effective Size, Mean (SD) | 2.97 (1.62) | 3.38 (1.65) | **<.001** |
| Max Degree, Mean (SD) | 4.38 (2.07) | 4.54 (1.99) | .055 |
| Mean Degree, Mean (SD) | 3.48 (1.77) | 3.43 (1.59) | .485 |
| *Network Composition* |  |  |  |
| Percent Kin, Mean (SD) | 55.89 (29.50) | 50.71 (28.84) | **<.001** |
| Standard Deviation of Age, Mean (SD) | 12.65 (6.80) | 11.96 (5.82) | .033 |
| Diversity of Sex, Mean (SD) | 67.26 (39.72) | 70.80 (30.26) | .016 |
| Diversity of Race, Mean (SD) | 6.00 (16.42) | 7.32 (17.03) | .061 |
| Percent contacted weekly or less, Mean (SD) | 15.89 (20.90) | 17.67 (21.15) | .043 |
| Percent known for less than 6 years, Mean (SD) | 11.81 (19.61) | 17.57 (23.65) | **<.001** |
| Percent who live over 15 miles away, Mean (SD) | 32.45 (27.82) | 38.93 (26.63) | **<.001** |
| Percent who drink, Mean (SD) | 12.43 (24.03) | 16.63 (26.55) | **<.001** |
| Percent who smoke, Mean (SD) | 8.87 (18.14) | 7.17 (15.85) | .017 |
| Percent non exercisers, Mean (SD) | 35.28 (31.60) | 39.06 (30.43) | .004 |
| Percent bad diet, Mean (SD) | 23.04 (27.61) | 22.75 (28.08) | .806 |
| Percent with a negative health influence, Mean (SD) | 36.16 (30.43) | 36.34 (30.16) | .890 |

- a. Bolded p values meet the significance threshold of $p < .002$ ( $\alpha = 0.05$ , corrected for 18 comparisons).

Next, we examined the association between each structural and compositional network feature in relation to PDDS, MSRS-R and PROMIS Physical Function in pwMS during the COVID-19 pandemic, after adjusting for age, disease duration, employment, and income in linear regression models (**Table 3**). PwMS who had a higher percent of network members with a perceived negative health influence had higher MSRS-R scores, indicating greater MS symptom burden and disability (Beta[95% CI]=2.181[1.082, 3.279], p<.001). Interestingly, none of the specific network health behavior features (*e.g.,* smoking, alcohol use, bad diet, and sedentary lifestyle) that might be construed as a negative health influence showed statistically significant association with MSRS-R. No other network features had significant association with any of the PROs after Bonferroni correction.

**Table 3.** Cross-sectional analysis of personal social network features in relation to patient-reported outcomes in people with multiple sclerosis during the COVID-19 pandemic.

|  | PDDS <sup>a</sup> |  |  |  |  | MSRS-R <sup>b</sup> |  |  |  |  | PROMIS <sup>c</sup> |  |  |  |  |
| --- | --- | --- | --- | --- | --- | --- | --- | --- | --- | --- | --- | --- | --- | --- | --- |
|  | N <sup>f</sup> | Beta <sup>d</sup> | 95% CI (lower) | 95% CI (upper) | p value <sup>e</sup> | N <sup>f</sup> | Beta <sup>d</sup> | 95% CI (lower) | 95% CI (upper) | p value <sup>e</sup> | N <sup>f</sup> | Beta <sup>d</sup> | 95% CI (lower) | 95% CI (upper) | p value <sup>e</sup> |
| *Network Structure* |  |  |  |  |  |  |  |  |  |  |  |  |  |  |  |
| Size | 737 | -0.017 | -0.05 | 0.016 | .314 | 737 | -0.032 | -0.117 | 0.053 | .462 | 728 | 0.165 | 0.001 | 0.33 | .049 |
| Density | 704 | -0.051 | -0.569 | 0.467 | .848 | 704 | 0.115 | -1.212 | 1.443 | .865 | 695 | -0.369 | -2.933 | 2.194 | .777 |
| Constraint | 704 | -0.001 | -0.009 | 0.006 | .681 | 704 | -0.004 | -0.023 | 0.014 | .627 | 695 | -0.006 | -0.041 | 0.029 | .753 |
| Effective Size | 704 | 0.006 | -0.079 | 0.091 | .895 | 704 | -0.044 | -0.262 | 0.173 | .689 | 695 | 0.15 | -0.27 | 0.569 | .484 |
| Maximum Degree | 704 | 0.018 | -0.05 | 0.085 | .603 | 704 | 0.02 | -0.153 | 0.192 | .821 | 695 | 0.017 | -0.316 | 0.35 | .920 |
| Mean Degree | 704 | 0.012 | -0.066 | 0.089 | .767 | 704 | 0.078 | -0.121 | 0.277 | .441 | 695 | -0.049 | -0.433 | 0.335 | .802 |
| *Network Composition* |  |  |  |  |  |  |  |  |  |  |  |  |  |  |  |
| Percent Kin | 713 | 0.191 | -0.25 | 0.632 | .395 | 713 | 0.614 | -0.514 | 1.742 | .285 | 704 | -1.356 | -3.541 | 0.83 | .224 |
| Standard Deviation of Age | 534 | -0.01 | -0.032 | 0.012 | .362 | 534 | -0.013 | -0.071 | 0.045 | .656 | 526 | 0.025 | -0.082 | 0.132 | .645 |
| Diversity of Sex | 707 | 0.148 | -0.277 | 0.574 | .493 | 707 | 0.24 | -0.844 | 1.324 | .664 | 698 | -1.143 | -3.249 | 0.962 | .287 |
| Diversity of Race | 701 | 0.545 | -0.403 | 1.492 | .260 | 701 | -1.058 | -3.48 | 1.365 | .392 | 692 | -1.506 | -6.195 | 3.183 | .529 |
| Percent contacted weekly or less | 713 | 0.394 | -0.245 | 1.034 | .226 | 713 | -0.601 | -2.238 | 1.036 | .471 | 704 | -1.13 | -4.306 | 2.046 | .485 |
| Percent known for less than 6 years | 713 | -0.215 | -0.909 | 0.478 | .542 | 713 | -0.05 | -1.825 | 1.725 | .956 | 704 | 1.326 | -2.11 | 4.763 | .449 |
| Percent who live over 15 miles away | 713 | -0.199 | -0.667 | 0.269 | .404 | 713 | 0.176 | -1.023 | 1.374 | .774 | 704 | 2.306 | -0.008 | 4.619 | .051 |
| Percent who drink | 713 | -0.326 | -0.92 | 0.268 | .282 | 713 | -0.391 | -1.912 | 1.13 | .614 | 704 | 2.979 | -0.003 | 5.96 | .050 |
| Percent who smoke | 713 | -0.139 | -0.882 | 0.603 | .712 | 713 | 1.046 | -0.853 | 2.944 | .280 | 704 | -2.155 | -5.827 | 1.518 | .250 |
| Percent non exerciser s | 713 | -0.069 | -0.486 | 0.348 | .746 | 713 | 0.771 | -0.295 | 1.837 | .156 | 704 | -0.281 | -2.351 | 1.788 | .790 |
| Percent bad diet | 713 | 0.048 | -0.437 | 0.534 | .845 | 713 | 1.242 | 0.003 | 2.481 | .049 | 704 | -2.794 | -5.189 | -0.4 | .022 |
| Percent with a negative health influence | 713 | -0.281 | -0.714 | 0.153 | .204 | 713 | 2.181 | 1.082 | 3.279 | **<.001** | 704 | -0.65 | -2.795 | 1.494 | .552 |

- a. PDDS: Patient-Determined Disease Steps. - b. MSRS-R: Multiple Sclerosis Rating Scale-Revised. - c. PROMIS: Patient-Reported Outcomes Measurement Information System Physical Function. - d. Adjusted for potential confounders (as identified in eFigure-1) including age, disease duration, employment, and income. - e. Bolded p values met the significance threshold of $p < .00092$ ( $\alpha = 0.05$ , corrected for 54 comparisons). - f. The smaller sample size in this analysis than the overall cohort size was due to the requirement for participants who had complete data elements for network feature, neurological outcome, and *all* covariates.

For comparison, we examined the association between each network feature with PROMIS Physical Function during the COVID-19 pandemic in controls (**eTable 2**). We adjusted for age, employment, and income, but disease duration was not applicable for controls. We found that lower maximum degree (Beta[95% CI]=0.383[0.141, 0.626], p=.002), higher percent who smoke (Beta[95% CI]=-4.846[-7.919, -1.774], p=.002), and higher percent of people with a perceived negative health influence (Beta[95% CI]=-5.707[-7.405, -4.010], p<.001) were associated with worse PROMIS Physical Function scores in controls. Given that the percent of network members with a perceived negative health influence was associated with MSRS-R in pwMS and with PROMIS Physical Function in controls, respectively, this important compositional network feature that summarizes the health behaviors of the personal social network might contribute to the physical impairment in both pwMS and control participants.

### Moderation Analysis Assessing the Role of MS Diagnosis

We performed a moderation analysis to examine whether MS diagnosis influences the strength and direction of the association between network features and PROMIS physical function during the COVID-19 pandemic (**eTable 4, Figure 3**). In this joint analysis combining pwMS and controls, MS diagnosis moderated the direction and/or strength of the association between several network features (diversity of race, percent who live over 15 miles away, percent who drink, and percent with a perceived negative health influence) and PROMIS physical function. For diversity of race, the moderating effect of MS diagnosis was marginally significant (Beta[95% CI]=-5.878[-10.889, -0.868], p=.022, **eTable 4**) such that this network feature was not significantly associated with physical function in either pwMS (Slope[95% CI]=-2.968 [-6.893, 0.956], p=.138) or control participants (Slope[95% CI]=2.910 [-0.224, 6.044], p=.068) (**Figure 3B, eTable 5**). For percent who live over 15 miles away, MS diagnosis moderated the strength of association such that this network feature was significantly associated with worse physical function in pwMS (Slope[95% CI]=3.890[1.852, 5.928], p<.001), but not in controls (Slope[95% CI]=-0.772[-2.765, 1.221], p=.448) (**eTable 5, Figure 3C**). Similarly, percent (of network members) who drink was associated with worse physical function in pwMS (Slope[95% CI]=3.716[1.392, 6.039], p=.002), but again not in controls (Slope[95% CI]=-0.982[-2.966, 1.003], p=.332) (**eTable 5, Figure 3D**). For percent with a perceived negative health influence, while both groups exhibited the same direction of association with physical function, pwMS had smaller magnitude of the association (Slope[95% CI]=-2.174 [-4.019, -0.329], p=.021) than controls (Slope[95% CI]=-6.601[-8.494, -4.708], p<.001) (**eTable 5, Figure 3E**).

### Longitudinal Analysis Comparing the Pandemic Period with the Pre-pandemic Baseline

We conducted a longitudinal analysis using a subset of pwMS and control participants who completed the PERSNET both preceding and during the COVID-19 pandemic. During the pandemic, both pwMS and control participants reported lower percent of people contacted weekly than the pre-pandemic baseline (p<.001), reflecting the widespread social isolation during the COVID-19 pandemic. When compared to the pre-pandemic baseline, both pwMS and controls had a reduction in network size (pwMS p=.003, control p<.001), effective size (pwMS p=.007, control p=.013), maximum degree (pwMS p=.01; control p<.001), and percent contacted weekly or less (pwMS p<.001; control p<.001) as well as an increase in constraint (pwMS p=.006, control p=.001) (**Table 4**). This indicates personal social network contraction for both pwMS and controls during the pandemic period. There was an increase in the percentage of kin (increase from 46.06% to 56.36%, p=.003) in the networks of pwMS during the COVID-19 pandemic, which was not seen in control participants.

Finally, we examined whether changes in network features due to the pandemic (*i.e.,* pandemic values minus the most recent pre-pandemic baseline) in pwMS were associated with the PROs during the COVID-19 pandemic (**eTable 3**, **Figure 2**). We found no significant association between changes in network features and pandemic PROs. As a confirmation of these findings, there was no difference between the observed and expected distribution of p-values of association between changes in network features and pandemic PROs in the permuted omnibus test (PDDS p=.880, MSRS-R p=.287, PROMIS p=.278).

**Figure 2.**
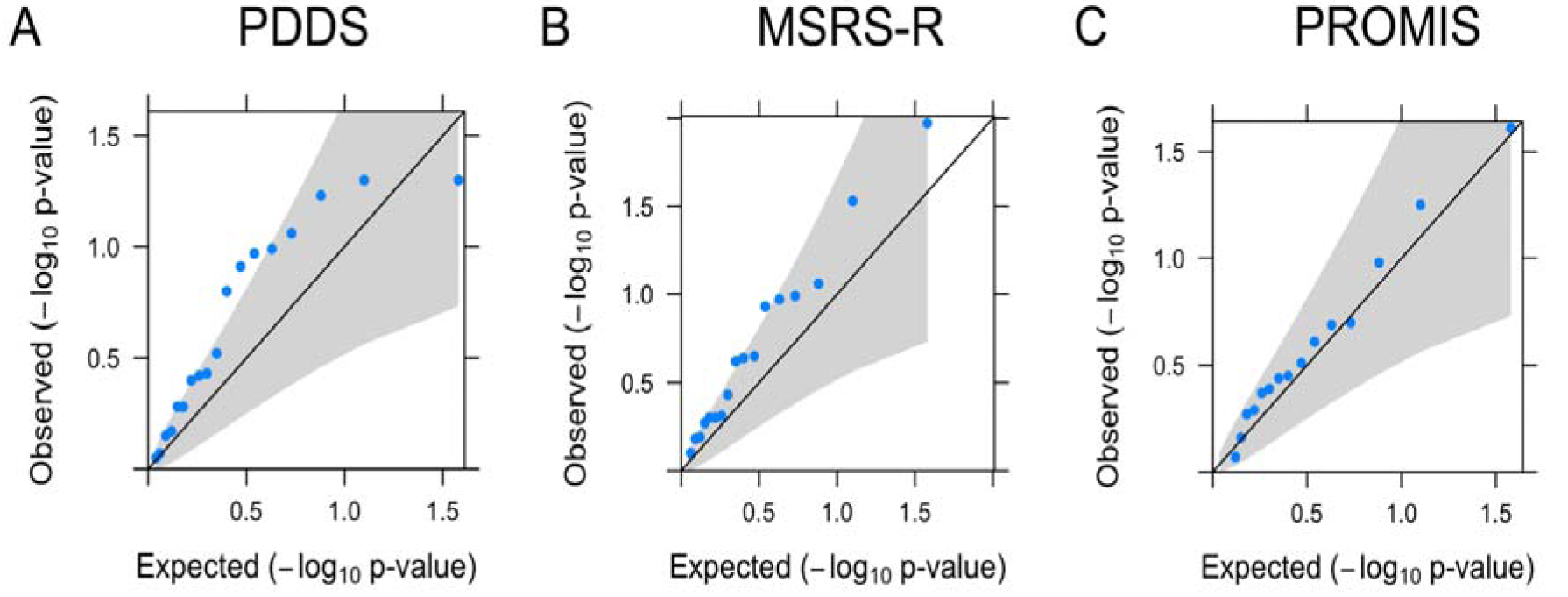
Quantile-quantile plots demonstrating longitudinal changes in personal social network features (due to the COVID-19 pandemic) in relation to patient-reported outcomes during the COVID-19 pandemic in people with multiple sclerosis. Comparison of observed and expected associations between the *difference* in each of quantitative structural and compositional personal social network features (pandemic value minus the pre-pandemic baseline) in relation to the three interrelated measures of neurological functions (A: Patient-Determined Disease Steps [PDDS]; B: Multiple Sclerosis Rating Scale-Revised [MSRS-R]; C: Patient-Reported Outcomes Measurement Information System [PROMIS] Physical Function) during the pandemic in people with multiple sclerosis after adjusting for age, disease duration, employment, income, study cohort, and time lapse between pre-pandemic and pandemic PERSNET assessment. Expected p values (-log10[p-value]) were plotted on the x-axis and observed p values (-log10[p-value]) on the y-axis. The grey area encompasses the 95% confidence intervals. Points outside the grey area were considered statistically significant without adjustment for multiple comparisons.

**Figure 3.**
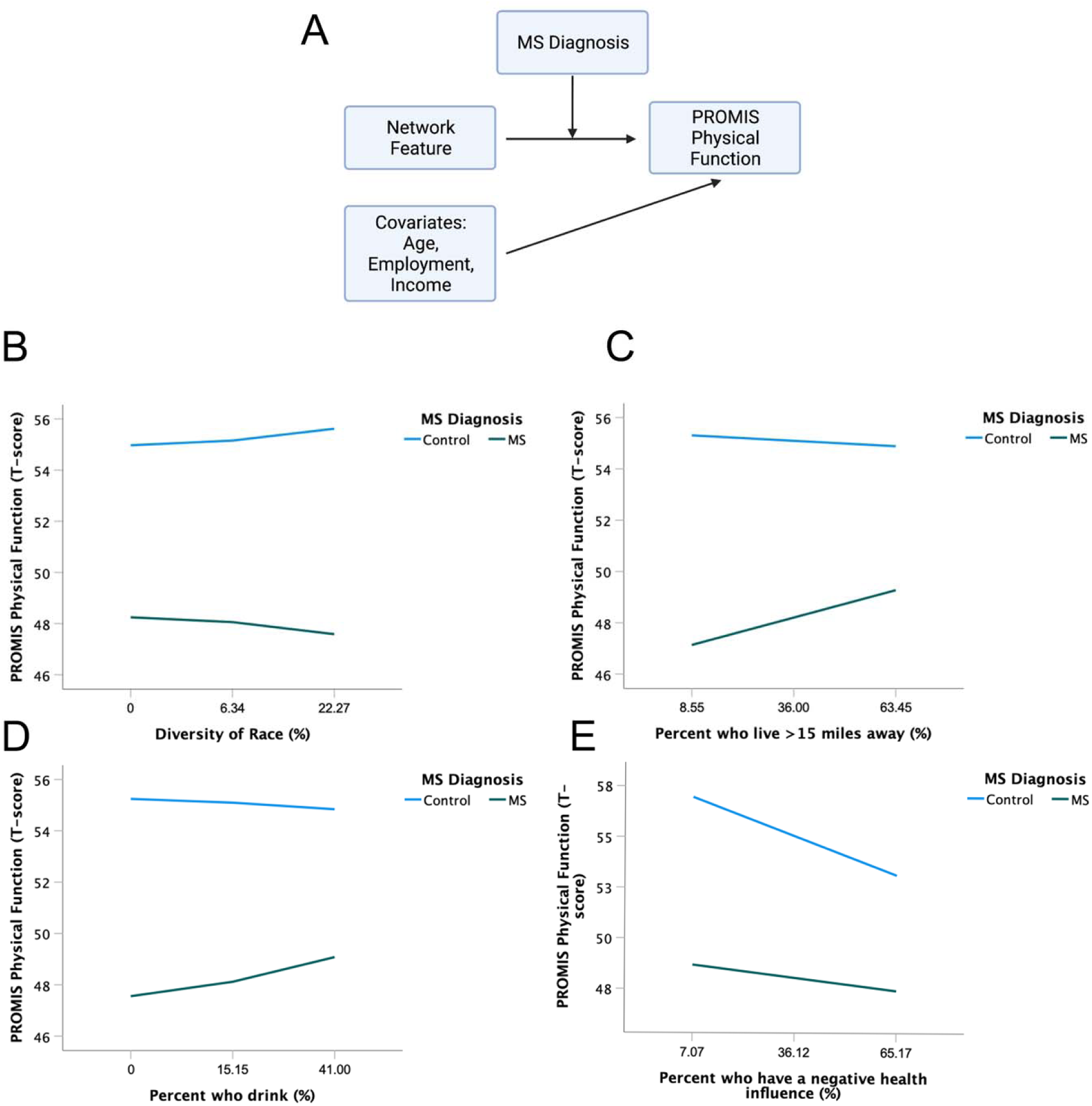
Role of multiple sclerosis diagnosis in moderating the association between personal social network features and PROMIS Physical Function in people with multiple sclerosis and control participants. A: In moderation analysis, we examined the association between each network feature and PROMIS Physical Function after combining the data from people with multiple sclerosis and controls and after adjusting for age, employment and income as covariates and further investigated whether the multiple sclerosis (MS) diagnosis moderated the direction or strength of this association. We found that MS diagnosis moderated the direction of the association between diversity of race (B), percent who live >15 miles away (C), percent who drink (D), and the strength of association between percent with a perceived negative health influence (E) and PROMIS Physical Function. Values on x-axis indicate the 25th, 50th, 75th percentile.

**Table 4.** Comparison of personal social network features in people with multiple sclerosis and control participants during the COVID-19 pandemic as compared to the within-subject pre-pandemic baseline.

|  | Control:<br>Pre-Pandemic<br>Baseline | Control:<br>During<br>Pandemic | p value <sup>a</sup> | MS:<br>Pre-Pandemic<br>Baseline | MS:<br>During<br>Pandemic | p value <sup>a</sup> |
| --- | --- | --- | --- | --- | --- | --- |
| N | 136 | 136 |  | 230 | 230 |  |
| ***Network Structure*** |  |  |  |  |  |  |
| Network Size,<br>Mean (SD) | 8.18 (4.05) | 6.44 (3.92) | **<.001** | 8.02 (5.70) | 6.63 (4.16) | **.003** |
| Density, Mean<br>(SD) | 0.69 (0.24) | 0.72 (0.24) | .239 | 0.74 (0.25) | 0.77 (0.24) | .137 |
| Constraint,<br>Mean (SD) | 48.07 (13.36) | 53.99 (16.31) | **.001** | 52.24 (15.81) | 56.77 (18.91) | **.006** |
| Effective<br>Size, Mean (SD) | 3.85 (1.56) | 3.40 (1.55) | **.013** | 3.30 (1.59) | 2.90 (1.50) | **.007** |
| Max Degree,<br>Mean (SD) | 5.38 (1.94) | 4.55 (2.06) | **<.001** | 4.78 (1.86) | 4.32 (1.92) | **.010** |
| Mean Degree,<br>Mean (SD) | 3.95 (1.71) | 3.47 (1.72) | **.018** | 3.70 (1.64) | 3.47 (1.66) | .142 |
| ***Network Composition*** |  |  |  |  |  |  |
| Percent Kin,<br>Mean (SD) | 47.69 (22.86) | 50.27 (26.87) | .378 | 46.06 (29.34) | 54.36 (30.16) | **.003** |
| Standard<br>Deviation of Age,<br>Mean (SD) | 12.67 (5.20) | 11.60 (6.26) | .153 | 12.69 (6.05) | 13.74 (6.69) | .225 |
| Diversity of<br>Sex, Mean (SD) | 77.61 (24.00) | 70.31 (30.73) | **.026** | 57.14 (39.82) | 52.17 (57.90) | .292 |
| Diversity of<br>Race, Mean (SD) | 7.97 (18.55) | 8.31 (18.43) | .874 | 5.66 (15.54) | 7.13 (20.47) | .392 |
| Percent<br>contacted weekly or<br>less, Mean (SD) | 30.34 (26.35) | 18.78 (22.23) | **<.001** | 23.48 (24.95) | 14.89 (20.95) | **<.001** |
| Percent known<br>for less than 6 years,<br>Mean (SD) | 21.22 (23.58) | 18.31 (21.95) | .270 | 13.26 (19.83) | 10.41 (18.39) | .115 |
| Percent who<br>live over 15 miles<br>away, Mean (SD) | 41.37 (28.04) | 38.88 (28.42) | .450 | 31.48 (25.57) | 33.76 (29.36) | .379 |
| Percent who<br>drink, Mean (SD) | 17.85 (27.04) | 19.13 (27.58) | .704 | 8.85 (18.87) | 10.32 (21.66) | .512 |
| Percent who<br>smoke, Mean (SD) | 12.70 (24.28) | 8.09 (18.57) | .064 | 12.72 (22.43) | 9.13 (17.90) | .061 |
| Percent non<br>exercisers, Mean<br>(SD) | 41.93 (27.91) | 39.86 (30.42) | .545 | 39.33 (30.94) | 31.88 (33.13) | **.014** |
| Percent bad<br>diet, Mean (SD) | 31.32 (28.73) | 25.74 (27.77) | .108 | 24.95 (29.57) | 24.87 (30.21) | .982 |
| Percent who<br>have a negative<br>health influence, Mean<br>(SD) | 36.29 (24.75) | 35.59 (25.14) | .819 | 36.77 (29.71) | 38.02 (28.61) | .697 |

a. Bolded p values meet the significance threshold of $p < .05$ .

## DISCUSSION

Our quantitative assessment of the personal social network environment of pwMS during the COVID-19 pandemic has several key findings. First, a higher percent of network members with a perceived negative health influence was associated with both greater neurological symptom disability in pwMS and worse physical function in controls during the pandemic, validating pre-pandemic findings and suggesting a shared contribution of this important feature (which summarizes the health behaviors of an individual’s personal social network) towards physical function in pwMS and controls. Second, personal social networks of pwMS and control participants experienced contraction during the COVID-19 pandemic when compared to the pre-pandemic baseline and the personal social networks of pwMS comprised of higher percent kin than control participants during the pandemic. Finally, changes in personal social network features related to the COVID-19 pandemic (when compared to the pre-pandemic baseline) in pwMS were not associated with worsening disability during the pandemic, suggesting an element of neurological resilience despite the perturbation in social environment and connections.

Our study design has several novel aspects. First, this is the first known direct comparison of the personal social networks of pwMS and control participants during the height of the COVID-19 pandemic when public health measures enacted to reduce contagion were widespread. Second, this is the first effort to longitudinally quantify changes in personal social networks of pwMS and control participants due to the COVID-19 pandemic. Third, this study evaluated the differential impact of MS diagnosis on the associations between personal network features and clinical outcomes in terms of both strength and directionality. Finally, this largest cross-sectionally and longitudinal quantitative examination of the association of personal social networks in relation to neurological and physical functions (not just in MS) explored the potential of neurological resilience secondary to social perturbation in the setting of the COVID-19 pandemic.

During the pandemic, the personal social networks of pwMS and controls both contracted when compared to the pre-pandemic period, as indicated by the increased constraint as well as decreased network size, effective size and maximum degree. The percentage of people contacted weekly or less also decreased for both pwMS and controls, suggesting increased social isolation. Notably, the personal social networks of pwMS were already more tightly knit during the pre-pandemic period than control participants, likely due to the vulnerability of the personal social networks of pwMS as reported in our previous studies.^8, 9^ The increase in percentage of kin in the personal social networks of pwMS during the pandemic was not seen in control participants. This could potentially be attributable to the differential impact of social isolation policies enacted during the pandemic on pwMS.^15–17^

The finding of the association between a higher percent of network members with a perceived negative health influence and worse neurological disability in pwMS and worse physical function in controls during the COVID-19 pandemic validated our prior findings from the pre-pandemic period.^8^ These findings are unlikely to be spurious given the relatively high proportion of participants (both pwMS and controls) whose network members have a perceived negative health influence. This compositional network feature (*i.e.,* percent of network members with a perceived negative health influence) likely better captured the overall negative health influence within an individual’s personal social network than other compositional features denoting a specific negative health behavior among the network members. We hypothesize that perceived negative health influences in one’s social network may decrease an individual’s likelihood of engaging in healthy behaviors (*e.g.,* moderate alcohol consumption, abstention from smoking, regular exercise, healthy diet, medication adherence) that may reduce overall comorbidities and MS-related disease activity as well as disability accumulation. We could test the causality in future intervention studies. For example, identifying persons with vulnerable network characteristics such as high percent of negative influence in the personal social network could help clinicians to provide closer monitoring of medication adherence, more concrete behavioral changes to reduce toxic exposures, and more encouragement to promote healthy behaviors, particularly in scenarios of perturbation to the social environment.

The previously reported association between tightly knit personal social networks and worse physical function was not observed in this study.^9^ We postulate several explanations. First, the contraction of social networks evidenced by a decrease in network size, decrease in effective size, increase in constraint, decrease in maximum degree, and decrease in people contacted weekly or less in pwMS as well as controls during the pandemic as compared to the pre-pandemic baseline are likely attributable to certain public health measures aiming to prevent the spread of COVID-19 given the data collection period of this study. Personal social networks broadly contracted and became more tight-knit among pwMS during the COVID-19 pandemic, whereas there was greater diversity (*e.g.,* higher effective network size, lower constraint) in the personal networks of pwMS before the pandemic. In our prior cross-sectional study, the direction of the association between network features and physical function could not be fully determined.^9^ It is possible that pwMS with better neurological and physical function might have more diverse personal social networks prior to the COVID-19 pandemic, but the overall contraction of social networks during the pandemic among pwMS might have masked the association between tightly knit networks and worse physical function. Future longitudinal analysis examining the change in network features from the pandemic period to the post-pandemic period may provide insight if the network diversity reverts to the pre-pandemic baseline versus if the new baseline persists. Second, since the contraction of personal social networks during the COVID-19 pandemic was a relatively recent and sudden event, there may not have been sufficient time for the onset of observable changes in neurological function. Assessment of neurological function at future time points could inform the long-term impact of social network changes due to the pandemic. Finally, the lack of association between longitudinal changes in social network features (comparing the pandemic period with the pre-pandemic period) and PROs suggests the possibility of neurological resilience in pwMS despite the adverse impact of the pandemic on the social fabric. In pwMS, psychological resilience has been shown to be associated with better physical outcomes.^35, 36^ PwMS have tightly knit personal social networks comprised a high proportion of kin whose support could potentially help preserve neurological function in pwMS (*e.g.,* family members help patients get to clinic or rehabilitation visits). Future studies exploring the relationships between personal social networks, social support, and neurological resilience would confirm this hypothesis.

The moderation analysis examined the interacting effect of MS diagnosis on the association between personal social network and PROs in a joint analysis of pwMS and controls. MS diagnosis moderated the association between specific personal social network features and physical function with respect to the direction (*i.e.,* diversity of race, percent of people who live over 15 miles away, percent of people who drink) or strength (*i.e.,* percent of people with a perceived negative health influence) of the associations. Some of the difference in results between the moderation analysis and the cross-sectional analysis for pwMS could be due to the inclusion of MS diagnosis as a moderator variable (affecting the strength and/or direction of association between network features and PROs) and inability to adjust for disease duration in this joint analysis as this covariate is not applicable for controls. Notably, lower percent of people who live over 15 miles away and lower percent of people who drink within the personal social networks of pwMS (but not controls) were each associated with worse physical function. PwMS with low physical function may need more support, which may explain their personal social networks comprising low percent of people who live far away and high proportion of kin who are more likely to live in the same household or in the vicinity. PwMS with low physical function may also be more inclined to seek out individuals whose healthy behaviors (*e.g.,* people who less likely to consume alcohol) could have a positive health influence. Reassuringly, the association between a higher percent of people with a perceived negative health influence and worse physical function in pwMS (and control participants) persisted. These consistent findings suggest that public health interventions broadly expanding the personal social networks of pwMS and reducing the negative health behaviors of personal social network members might improve health outcomes of all members, *i.e.,* beneficial contagion.

This study has several strengths. First, the longitudinal analysis of changes in personal social networks due to the COVID-19 pandemic in comparison to the pre-pandemic baseline had a within-subjects design. Consequently, we postulated that changes in network features suggestive of personal network contraction in the same participants during the COVID-19 pandemic (when compared to their pre-pandemic baseline) is likely attributable to the pandemic, possibly due to the necessary public health measures. Second, we used three independent but interrelated PROs as a pragmatic method to assess real-world neurological disability and physical function during the early pandemic when clinical research participation became severely restricted. These validated PROs, two specifically for MS and one generalizable across health and disease, have shown strong correlations with physician-rated measures of neurological function (*e.g.,* Expanded Disability Status Scale) in prior studies.^26–30^ Third, we conducted both cross-sectional and longitudinal analyses in not only pwMS but also control participants, which enabled comparison of the differential impact of the pandemic on the personal social networks. Fourth, the moderation analysis examining the role of MS diagnosis on the association between network features and physical function enabled exploration of the strength and directionality of these complex relationships. Finally, we leveraged a large multi-center dataset representative of the Northeastern and Mid-Atlantic regions of the United States with greater geographic diversity than prior studies, potentially increasing the generalizability of the study findings.

Our study also has limitations. First, the sample size of the longitudinal analysis was limited by the relatively modest number of participants with quantified pre-pandemic personal social networks as baseline. As such, the study did not have sufficient power for subgroup analysis stratified by demographic or clinical subtypes. Second, we sampled participants for their personal networks and neurological outcomes at one time point during the COVID-19 pandemic. The COVID-19 cases varied across areas and throughout the pandemic, while study participants resided in urban, suburban, and rural environments that were differentially impacted during the pandemic. Nevertheless, most of the study populations shared broader geographic regions (Northeastern and Mid-Atlantic USA) and completed the study response during the early period of the pandemic when federal and state level mandates were relatively uniform in terms of lockdown, social distancing, and other mitigation guidelines.

In conclusion, this study highlights the impact of the COVID-19 pandemic on personal social networks in pwMS and control participants and suggests potential resilience despite the social perturbation. Our findings generate important hypotheses for testing future interventions that may modify personal social networks to broadly improve neurological outcomes. Future longitudinal studies examining the long-term impact of the COVID-19 pandemic on the evolution of personal social networks and neurological outcomes in people with chronic neurological disorders such as MS are warranted.

## Data Availability

The code for this project is available at https://github.com/shruthi1094/pnq. De-identified data are available upon request to the corresponding author and with permission from each participating center.

## SUPPLEMENTAL MATERIAL

**eFigure 1.**
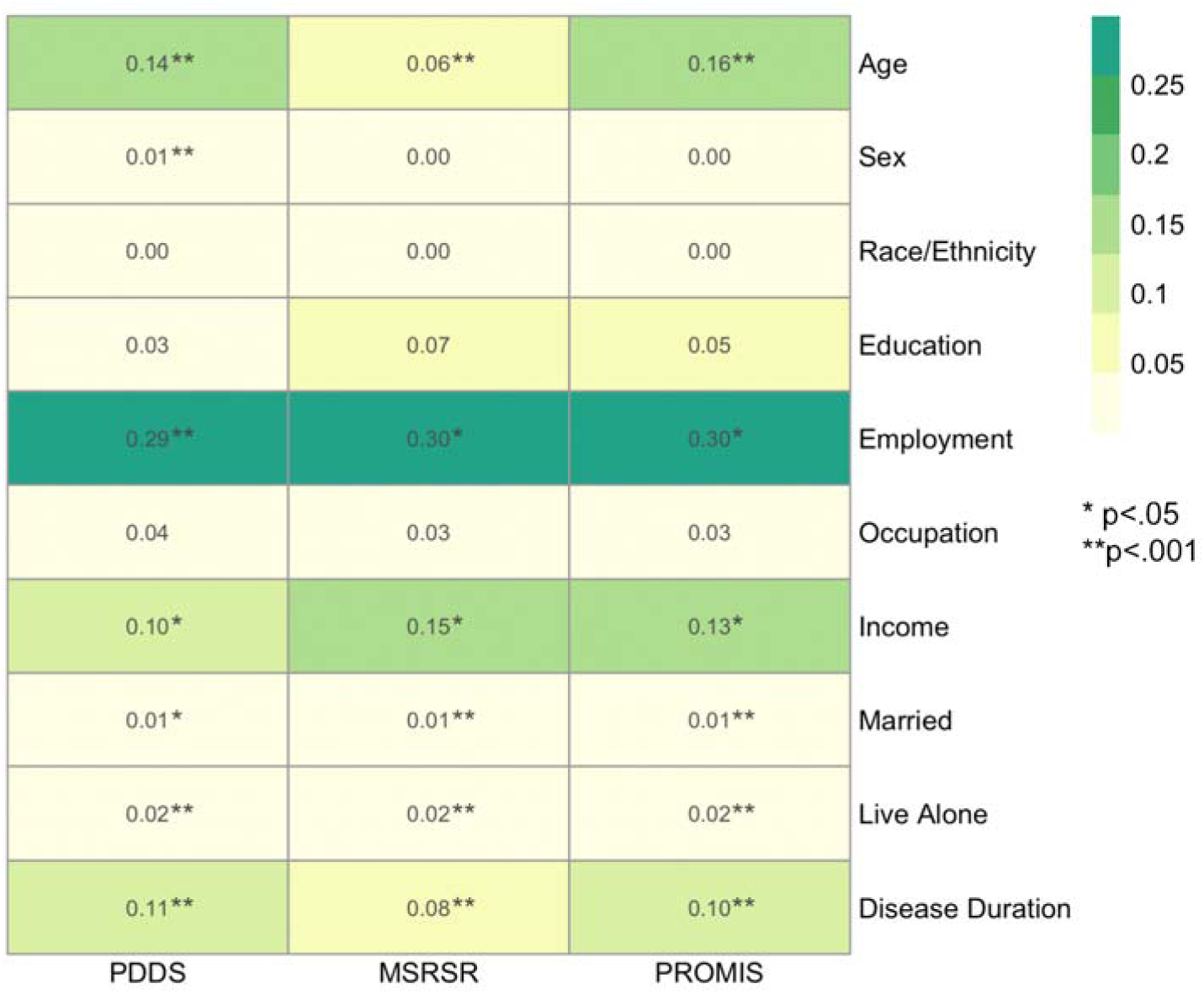
Correlation between potential confounders and patient-reported outcomes. Using a data-driven approach for covariate adjustment, we calculated the Pearson correlation coefficients (R^2^, actual value and heatmap intensity shown inside each box) and the p-values (marked as meeting the pre-defined significance threshold) of potential confounding variables in relation to patient-reported outcomes. Covariates were selected for adjustment in downstream analysis if over 70% of participants had the pertinent information (frequency > 70%), the average correlation coefficient across all patient-reported outcomes was over 0.1 (R^2^ > 0.1), and the p-values met the nominal significance threshold (p<.05).

**eTable 1.** Participant characteristics of cross-sectional cohorts by recruitment site.

|  | Overall:<br>MS | Overall:<br>Control | MS | Control | MS | Control | MS | Control | MS | Control | MS | Control | MS | Control | MS | Control | MS | Control |
| --- | --- | --- | --- | --- | --- | --- | --- | --- | --- | --- | --- | --- | --- | --- | --- | --- | --- | --- |
|  |  |  | Cohort<br>1<sup>a</sup> | Cohort<br>1<sup>a</sup> | Cohort<br>2<sup>b</sup> | Cohort<br>2<sup>b</sup> | Cohort<br>3<sup>c</sup> | Cohort<br>3<sup>c</sup> | Cohort<br>4<sup>d</sup> | Cohort<br>4<sup>d</sup> | Cohort<br>5<sup>e</sup> | Cohort<br>5<sup>e</sup> | Cohort<br>6<sup>f</sup> | Cohort<br>6<sup>f</sup> | Cohort<br>7<sup>g</sup> | Cohort<br>7<sup>g</sup> | Cohort<br>8<sup>h</sup> | Cohort<br>8<sup>h</sup> |
| N | 1130 | 1250 | 282 | 60 | 60 | 165 | 67 | 2 | 129 | 222 | 114 | 34 | 127 | 12 | 290 | 23 | 61 | 732 |
| Age, Mean (SD) | 50.7<br>(12.1) | 44.4<br>(12.1) | 50.8<br>(12.9) | 53.1<br>(13.7) | 49.3<br>(11.5) | 53.6<br>(17.2) | 46.7<br>(11.0) | 26.5<br>(38.9) | 49.7<br>(10.9) | 41.6<br>(9.1) | 48.5<br>(13.5) | 52.7<br>(17.1) | 55.4<br>(11.8) | 52.5<br>(20.1) | 51.6<br>(12.0) | 55.5<br>(15.1) | 48.1<br>(7.4) | 41.6<br>(8.3) |
| Sex, N (%) |  |  |  |  |  |  |  |  |  |  |  |  |  |  |  |  |  |  |
| Female | 925<br>(81.9) | 960<br>(76.8) | 229<br>(81.2) | 31<br>(51.7) | 52<br>(86.7) | 115<br>(69.7) | 51<br>(76.1) | 0<br>(0.0) | 104<br>(80.6) | 183<br>(82.4) | 92<br>(80.7) | 19<br>(55.9) | 101<br>(79.5) | 7<br>(58.3) | 242<br>(83.4) | 16<br>(69.6) | 54<br>(88.5) | 589<br>(80.5) |
| Male | 203<br>(18.0) | 286<br>(22.9) | 53<br>(18.8) | 28<br>(46.7) | 7<br>(11.7) | 48<br>(29.1) | 16<br>(23.9) | 2<br>(100.0) | 25<br>(19.4) | 39<br>(17.6) | 22<br>(19.3) | 15<br>(44.1) | 25<br>(19.7) | 5<br>(41.7) | 48<br>(16.6) | 7<br>(30.4) | 7<br>(11.5) | 142<br>(19.4) |
| Other | 2<br>(0.2) | 4<br>(0.3) | 0<br>(0.0) | 1<br>(1.7) | 1<br>(1.7) | 2<br>(1.2) | 0<br>(0.0) | 0<br>(0.0) | 0<br>(0.0) | 0<br>(0.0) | 0<br>(0.0) | 0<br>(0.0) | 1<br>(0.8) | 0<br>(0.0) | 0<br>(0.0) | 0<br>(0.0) | 0<br>(0.0) | 1<br>(0.1) |
| Race, N (%) |  |  |  |  |  |  |  |  |  |  |  |  |  |  |  |  |  |  |
| African or African American | 46<br>(4.1) | 12<br>(1.0) | 8<br>(2.8) | 0<br>(0.0) | 1<br>(1.7) | 1<br>(0.6) | 7<br>(10.4) | 0<br>(0.0) | 4<br>(3.1) | 2<br>(0.9) | 2<br>(1.7) | 1<br>(3.0) | 6<br>(4.7) | 0<br>(0.0) | 17<br>(6.0) | 0<br>(0.0) | 1<br>(1.6) | 8<br>(1.1) |
| American Indian or Alaskan Native or Native Hawaiian | 10<br>(0.9) | 6<br>(0.5) | 0<br>(0.0) | 0<br>(0.0) | 0<br>(0.0) | 1<br>(0.6) | 1<br>(1.5) | 0<br>(0.0) | 0<br>(0.0) | 3<br>(1.4) | 7<br>(6.0) | 0<br>(0.0) | 0<br>(0.0) | 0<br>(0.0) | 2<br>(0.7) | 0<br>(0.0) | 0<br>(0.0) | 2<br>(0.3) |
| Asian | 8<br>(0.7) | 8<br>(0.6) | 0<br>(0.0) | 0<br>(0.0) | 2<br>(3.3) | 2<br>(1.2) | 1<br>(1.5) | 0<br>(0.0) | 1<br>(0.8) | 0<br>(0.0) | 0<br>(0.0) | 2<br>(6.1) | 2<br>(1.6) | 0<br>(0.0) | 1<br>(0.4) | 0<br>(0.0) | 1<br>(1.6) | 4<br>(0.5) |
| White | 1043<br>(92.7) | 1208<br>(96.7) | 274<br>(97.2) | 60<br>(100.0) | 55<br>(91.7) | 160<br>(97.0) | 50<br>(74.6) | 2<br>(100.0) | 121<br>(93.8) | 213<br>(95.9) | 104<br>(89.7) | 30<br>(90.9) | 119<br>(93.7) | 12<br>(100.0) | 261<br>(92.2) | 23<br>(100.0) | 59<br>(96.7) | 708<br>(96.7) |
| Multi-racial | 9<br>(0.8) | 11<br>(0.9) | 0<br>(0.0) | 0<br>(0.0) | 1<br>(1.7) | 1<br>(0.6) | 5<br>(7.5) | 0<br>(0.0) | 0<br>(0.0) | 3<br>(1.4) | 3<br>(2.6) | 0<br>(0.0) | 0<br>(0.0) | 0<br>(0.0) | 0<br>(0.0) | 0<br>(0.0) | 0<br>(0.0) | 7<br>(1.0) |
| Other | 4<br>(0.4) | 3<br>(0.2) | 0<br>(0.0) | 0<br>(0.0) | 1<br>(1.7) | 0<br>(0.0) | 0<br>(0.0) | 0<br>(0.0) | 2<br>(1.6) | 0<br>(0.0) | 0<br>(0.0) | 0<br>(0.0) | 0<br>(0.0) | 0<br>(0.0) | 2<br>(0.7) | 0<br>(0.0) | 0<br>(0.0) | 1<br>(0.1) |
| Not Sure | 5<br>(0.4) | 1<br>(0.1) | 0<br>(0.0) | 0<br>(0.0) | 0<br>(0.0) | 0<br>(0.0) | 3<br>(4.5) | 0<br>(0.0) | 1<br>(0.8) | 1<br>(0.5) | 0<br>(0.0) | 0<br>(0.0) | 0<br>(0.0) | 0<br>(0.0) | 0<br>(0.0) | 0<br>(0.0) | 0<br>(0.0) | 2<br>(0.3) |
| Ethnicity, N (%) |  |  |  |  |  |  |  |  |  |  |  |  |  |  |  |  |  |  |
| Hispanic or Latino | 38<br>(3.4) | 31<br>(2.5) | 0<br>(0.0) | 0<br>(0.0) | 5<br>(8.3) | 2<br>(1.2) | 6<br>(9.0) | 0<br>(0.0) | 7<br>(5.4) | 6<br>(2.7) | 8<br>(6.9) | 1<br>(3.1) | 2<br>(1.6) | 0<br>(0.0) | 10<br>(3.5) | 0<br>(0.0) | 0<br>(0.0) | 22<br>(3.0) |
| Non- Hispanic | 1068<br>(94.9) | 1201<br>(96.3) | 282<br>(100.0) | 60<br>(100.0) | 54<br>(90.0) | 163<br>(98.8) | 55<br>(82.1) | 0<br>(0.0) | 122<br>(94.6) | 213<br>(95.9) | 103<br>(88.8) | 30<br>(93.8) | 122<br>(96.1) | 12<br>(100.0) | 269<br>(95.1) | 23<br>(100.0) | 61<br>(100.0) | 700<br>(95.8) |
| Not Sure | 19<br>(1.7) | 15<br>(1.2) | 0<br>(0.0) | 0<br>(0.0) | 1<br>(1.7) | 0<br>(0.0) | 6<br>(9.0) | 2<br>(100.0) | 0<br>(0.0) | 3<br>(1.4) | 5<br>(4.3) | 1<br>(3.1) | 3<br>(2.4) | 0<br>(0.0) | 4<br>(1.4) | 0<br>(0.0) | 0<br>(0.0) | 9<br>(1.2) |
| Education, N (%) |  |  |  |  |  |  |  |  |  |  |  |  |  |  |  |  |  |  |
| High school graduate | 70<br>(6.2) | 35<br>(2.8) | 28<br>(10.0) | 12<br>(20.0) | 4<br>(6.8) | 7<br>(4.2) | 1<br>(1.6) | 0<br>(0.0) | 6<br>(4.7) | 1<br>(0.5) | 7<br>(6.2) | 0<br>(0.0) | 7<br>(5.5) | 0<br>(0.0) | 16<br>(5.5) | 4<br>(17.4) | 1<br>(1.6) | 11<br>(1.5) |
| Some college | 154<br>(13.7) | 106<br>(8.5) | 47<br>(16.8) | 9<br>(15.0) | 4<br>(6.8) | 16<br>(9.7) | 4<br>(6.3) | 0<br>(0.0) | 18<br>(14.0) | 17<br>(7.7) | 24<br>(21.2) | 3<br>(8.8) | 21<br>(16.5) | 1<br>(8.3) | 31<br>(10.7) | 8<br>(34.8) | 5<br>(8.2) | 52<br>(7.1) |
| Associate degree | 111<br>(9.9) | 70<br>(5.6) | 43<br>(15.4) | 6<br>(10.0) | 6<br>(10.2) | 10<br>(6.1) | 3<br>(4.8) | 0<br>(0.0) | 14<br>(10.9) | 14<br>(6.3) | 8<br>(7.1) | 4<br>(11.8) | 20<br>(15.7) | 0<br>(0.0) | 13<br>(4.5) | 2<br>(8.7) | 4<br>(6.6) | 34<br>(4.7) |
| Bachelor's degree | 363<br>(32.4) | 441<br>(35.3) | 81<br>(28.9) | 16<br>(26.7) | 18<br>(30.5) | 61<br>(37.0) | 28<br>(44.4) | 1<br>(50.0) | 36<br>(27.9) | 76<br>(34.2) | 26<br>(23.0) | 8<br>(23.5) | 33<br>(26.0) | 6<br>(50.0) | 114<br>(39.3) | 3<br>(13.0) | 27<br>(44.3) | 270<br>(36.9) |
| Graduate degree | 418<br>(37.3) | 595<br>(47.6) | 81<br>(28.9) | 17<br>(28.3) | 27<br>(45.8) | 70<br>(42.4) | 27<br>(42.9) | 1<br>(50.0) | 54<br>(41.9) | 114<br>(51.4) | 48<br>(42.5) | 19<br>(55.9) | 44<br>(34.6) | 5<br>(41.7) | 114<br>(39.3) | 6<br>(26.1) | 23<br>(37.7) | 363<br>(49.7) |
| Employment, N (%) |  |  |  |  |  |  |  |  |  |  |  |  |  |  |  |  |  |  |
| Employed for wages | 544<br>(48.1) | 876<br>(70.1) | 121<br>(42.9) | 30<br>(50.0) | 33<br>(55.0) | 74<br>(44.8) | 36<br>(53.7) | 1<br>(50.0) | 66<br>(51.2) | 169<br>(76.1) | 60<br>(52.6) | 18<br>(52.9) | 47<br>(37.0) | 6<br>(50.0) | 140<br>(48.3) | 4<br>(17.4) | 41<br>(67.2) | 574<br>(78.4) |
| Homemaker | 63<br>(5.6) | 77<br>(6.2) | 15<br>(5.3) | 0<br>(0.0) | 1<br>(1.7) | 3<br>(1.8) | 4<br>(6.0) | 0<br>(0.0) | 5<br>(3.9) | 16<br>(7.2) | 8<br>(7.0) | 0<br>(0.0) | 7<br>(5.5) | 1<br>(8.3) | 21<br>(7.2) | 0<br>(0.0) | 2<br>(3.3) | 57<br>(7.8) |
| Military | 2<br>(0.2) | 0<br>(0.0) | 0<br>(0.0) | 0<br>(0.0) | 0<br>(0.0) | 0<br>(0.0) | 0<br>(0.0) | 0<br>(0.0) | 1<br>(0.8) | 0<br>(0.0) | 0<br>(0.0) | 0<br>(0.0) | 0<br>(0.0) | 0<br>(0.0) | 0<br>(0.0) | 0<br>(0.0) | 1<br>(1.6) | 0<br>(0.0) |
| Out of work and looking for work | 17<br>(1.5) | 27<br>(2.2) | 4<br>(1.4) | 0<br>(0.0) | 1<br>(1.7) | 4<br>(2.4) | 2<br>(3.0) | 0<br>(0.0) | 2<br>(1.6) | 8<br>(3.6) | 2<br>(1.8) | 1<br>(2.9) | 4<br>(3.1) | 0<br>(0.0) | 1<br>(0.3) | 0<br>(0.0) | 1<br>(1.6) | 14<br>(1.9) |
| Out of work but not currently looking for work | 29<br>(2.6) | 15<br>(1.2) | 7<br>(2.5) | 0<br>(0.0) | 2<br>(3.3) | 4<br>(2.4) | 1<br>(1.5) | 1<br>(50.0) | 5<br>(3.9) | 2<br>(0.9) | 4<br>(3.5) | 0<br>(0.0) | 6<br>(4.7) | 0<br>(0.0) | 4<br>(1.4) | 0<br>(0.0) | 0<br>(0.0) | 8<br>(1.1) |
| Retired | 187<br>(16.5) | 98<br>(7.8) | 46<br>(16.3) | 16<br>(26.7) | 6<br>(10.0) | 55<br>(33.3) | 8<br>(11.9) | 0<br>(0.0) | 20<br>(15.5) | 2<br>(0.9) | 18<br>(15.8) | 8<br>(23.5) | 30<br>(23.6) | 3<br>(25.0) | 56<br>(19.3) | 7<br>(30.4) | 3<br>(4.9) | 7<br>(1.0) |
| Self-employed | 63<br>(5.6) | 97<br>(7.8) | 11<br>(3.9) | 5<br>(8.3) | 2<br>(3.3) | 10<br>(6.1) | 3<br>(4.5) | 0<br>(0.0) | 7<br>(5.4) | 16<br>(7.2) | 7<br>(6.1) | 4<br>(11.8) | 5<br>(3.9) | 0<br>(0.0) | 25<br>(8.6) | 1<br>(4.3) | 3<br>(4.9) | 61<br>(8.3) |
| Student | 12<br>(1.1) | 18<br>(1.4) | 5<br>(1.8) | 0<br>(0.0) | 1<br>(1.7) | 8<br>(4.8) | 2<br>(3.0) | 0<br>(0.0) | 1<br>(0.8) | 4<br>(1.8) | 1<br>(0.9) | 0<br>(0.0) | 0<br>(0.0) | 0<br>(0.0) | 2<br>(0.7) | 0<br>(0.0) | 0<br>(0.0) | 6<br>(0.8) |
| Unable to work | 190<br>(16.8) | 39<br>(3.1) | 64<br>(22.7) | 9<br>(15.0) | 14<br>(23.3) | 5<br>(3.0) | 9<br>(13.4) | 0<br>(0.0) | 18<br>(14.0) | 5<br>(2.3) | 13<br>(11.4) | 3<br>(8.8) | 27<br>(21.3) | 2<br>(16.7) | 37<br>(12.8) | 11<br>(47.8) | 8<br>(13.1) | 4<br>(0.5) |
| Household Income (%) |  |  |  |  |  |  |  |  |  |  |  |  |  |  |  |  |  |  |
| 0 to \$19,999 | 74<br>(7.6) | 37<br>(3.1) | 32<br>(11.7) | 8<br>(13.6) | 6<br>(10.7) | 11<br>(6.9) | * | * | 6<br>(4.9) | 5<br>(2.4) | 6<br>(5.7) | 3<br>(8.8) | 7<br>(7.1) | 0<br>(0.0) | 12<br>(4.7) | 3<br>(20.0) | 5<br>(8.6) | 7<br>(1.0) |
| \$20,000 to \$34,999 | 85<br>(8.7) | 56<br>(4.7) | 26<br>(9.5) | 3<br>(5.1) | 6<br>(10.7) | 17<br>(10.6) | * | * | 10<br>(8.2) | 8<br>(3.9) | 12<br>(11.3) | 1<br>(2.9) | 15<br>(15.2) | 0<br>(0.0) | 13<br>(5.1) | 2<br>(13.3) | 3<br>(5.2) | 25<br>(3.5) |
| \$35,000 to \$49,999 | 97<br>(10.0) | 75<br>(6.3) | 43<br>(15.7) | 6<br>(10.2) | 8<br>(14.3) | 11<br>(6.9) | * | * | 8<br>(6.6) | 10<br>(4.8) | 8<br>(7.5) | 2<br>(5.9) | 9<br>(9.1) | 0<br>(0.0) | 16<br>(6.2) | 1<br>(6.7) | 5<br>(8.6) | 45<br>(6.4) |
| \$50,000 to \$64,999 | 92<br>(9.5) | 106<br>(8.9) | 28<br>(10.2) | 6<br>(10.2) | 5<br>(8.9) | 24<br>(15.0) | * | * | 9<br>(7.4) | 20<br>(9.7) | 12<br>(11.3) | 2<br>(5.9) | 15<br>(15.2) | 1<br>(12.5) | 21<br>(8.2) | 0<br>(0.0) | 2<br>(3.4) | 53<br>(7.5) |
| \$65,000 to \$79,999 | 79<br>(8.1) | 120<br>(10.1) | 26<br>(9.5) | 8<br>(13.6) | 4<br>(7.1) | 16<br>(10.0) | * | * | 14<br>(11.5) | 21<br>(10.1) | 10<br>(9.4) | 3<br>(8.8) | 8<br>(8.1) | 0<br>(0.0) | 12<br>(4.7) | 0<br>(0.0) | 5<br>(8.6) | 72<br>(10.2) |
| \$80,000 to \$94,999 | 76<br>(7.8) | 94<br>(7.9) | 15<br>(5.5) | 3<br>(5.1) | 2<br>(3.6) | 19<br>(11.9) | * | * | 11<br>(9.0) | 14<br>(6.8) | 6<br>(5.7) | 6<br>(17.6) | 11<br>(11.1) | 1<br>(12.5) | 26<br>(10.1) | 2<br>(13.3) | 5<br>(8.6) | 49<br>(6.9) |
| \$95,000 to \$109,999 | 80<br>(8.2) | 127<br>(10.7) | 29<br>(10.6) | 6<br>(10.2) | 2<br>(3.6) | 13<br>(8.1) | * | * | 14<br>(11.5) | 22<br>(10.6) | 4<br>(3.8) | 3<br>(8.8) | 11<br>(11.1) | 2<br>(25.0) | 15<br>(5.8) | 0<br>(0.0) | 5<br>(8.6) | 81<br>(11.5) |
| \$110,000 to \$124,999 | 85<br>(8.7) | 112<br>(9.4) | 26<br>(9.5) | 6<br>(10.2) | 4<br>(7.1) | 15<br>(9.4) | * | * | 7<br>(5.7) | 21<br>(10.1) | 12<br>(11.3) | 3<br>(8.8) | 3<br>(3.0) | 1<br>(12.5) | 26<br>(10.1) | 1<br>(6.7) | 7<br>(12.1) | 65<br>(9.2) |
| \$125,000 or higher | 303<br>(31.2) | 461<br>(38.8) | 49<br>(17.9) | 13<br>(22.0) | 19<br>(33.9) | 34<br>(21.2) | * | * | 43<br>(35.2) | 86<br>(41.5) | 35<br>(33.0) | 10<br>(29.4) | 20<br>(20.2) | 3<br>(37.5) | 116<br>(45.1) | 6<br>(40.0) | 21<br>(36.2) | 309<br>(43.8) |
| Married, N (%) | 786<br>(69.9) | 841<br>(67.4) | 185<br>(66.1) | 44<br>(73.3) | 37<br>(62.7) | 86<br>(52.1) | 45<br>(67.2) | 1<br>(50.0) | 95<br>(73.6) | 151<br>(68.0) | 84<br>(73.7) | 14<br>(42.4) | 82<br>(64.6) | 11<br>(91.7) | 214<br>(73.8) | 14<br>(60.9) | 44<br>(74.6) | 520<br>(71.2) |
| Live Alone, N (%) | 156<br>(14.8) | 186<br>(15.0) | 52<br>(18.5) | 11<br>(19.0) | 13<br>(22.0) | 43<br>(26.1) | * | * | 16<br>(12.6) | 27<br>(12.3) | 14<br>(12.3) | 8<br>(24.2) | 20<br>(15.9) | 1<br>(8.3) | 32<br>(11.0) | 6<br>(26.1) | 9<br>(15.3) | 90<br>(12.3) |
| Occupation, N (%) |  |  |  |  |  |  |  |  |  |  |  |  |  |  |  |  |  |  |
| Business owner | 35<br>(6.2) | 23<br>(2.4) | 9<br>(6.8) | 0<br>(0.0) | 2<br>(5.7) | 3<br>(3.6) | * | * | 5<br>(6.8) | 5<br>(2.7) | 2<br>(3.0) | 1<br>(4.5) | 3<br>(5.8) | 0<br>(0.0) | 11<br>(6.7) | 0<br>(0.0) | 3<br>(6.8) | 14<br>(2.2) |
| Executive, manager | 100<br>(17.6) | 157<br>(16.2) | 19<br>(14.4) | 5<br>(14.3) | 2<br>(5.7) | 5<br>(6.0) | * | * | 12<br>(16.4) | 31<br>(16.8) | 13<br>(19.4) | 8<br>(36.4) | 5<br>(9.6) | 1<br>(16.7) | 38<br>(23.0) | 2<br>(40.0) | 11<br>(25.0) | 105<br>(16.5) |
| Laborer, unskilled worker | 7<br>(1.2) | 4<br>(0.4) | 4<br>(3.0) | 1<br>(2.9) | 0<br>(0.0) | 0<br>(0.0) | * | * | 0<br>(0.0) | 0<br>(0.0) | 0<br>(0.0) | 0<br>(0.0) | 2<br>(3.8) | 0<br>(0.0) | 0<br>(0.0) | 0<br>(0.0) | 1<br>(2.3) | 3<br>(0.5) |
| Machine operator, inspector, bus/cab driver | 1<br>(0.2) | 0<br>(0.0) | 0<br>(0.0) | 0<br>(0.0) | 0<br>(0.0) | 0<br>(0.0) | * | * | 0<br>(0.0) | 0<br>(0.0) | 1<br>(1.5) | 0<br>(0.0) | 0<br>(0.0) | 0<br>(0.0) | 0<br>(0.0) | 0<br>(0.0) | 0<br>(0.0) | 0<br>(0.0) |
| Mechanic, electrician, skilled worker | 6<br>(1.1) | 14<br>(1.4) | 0<br>(0.0) | 0<br>(0.0) | 0<br>(0.0) | 1<br>(1.2) | * | * | 0<br>(0.0) | 3<br>(1.6) | 1<br>(1.5) | 0<br>(0.0) | 1<br>(1.9) | 0<br>(0.0) | 4<br>(2.4) | 0<br>(0.0) | 0<br>(0.0) | 10<br>(1.6) |
| Other | 80<br>(14.1) | 129<br>(13.3) | 19<br>(14.4) | 3<br>(8.6) | 5<br>(14.3) | 17<br>(20.2) | * | * | 11<br>(15.1) | 27<br>(14.6) | 12<br>(17.9) | 4<br>(18.2) | 6<br>(11.5) | 0<br>(0.0) | 21<br>(12.7) | 0<br>(0.0) | 6<br>(13.6) | 78<br>(12.3) |
| Professional | 282<br>(49.6) | 545<br>(56.1) | 66<br>(50.0) | 19<br>(54.3) | 22<br>(62.9) | 46<br>(54.8) | * | * | 38<br>(52.1) | 106<br>(57.3) | 33<br>(49.3) | 8<br>(36.4) | 28<br>(53.8) | 4<br>(66.7) | 73<br>(44.2) | 2<br>(40.0) | 22<br>(50.0) | 360<br>(56.7) |
| Sales or clerical worker | 54<br>(9.5) | 84<br>(8.6) | 14<br>(10.6) | 5<br>(14.3) | 4<br>(11.4) | 10<br>(11.9) | * | * | 5<br>(6.8) | 10<br>(5.4) | 5<br>(7.5) | 1<br>(4.5) | 7<br>(13.5) | 1<br>(16.7) | 18<br>(10.9) | 1<br>(20.0) | 1<br>(2.3) | 56<br>(8.8) |
| Service worker | 3<br>(0.5) | 16<br>(1.6) | 1<br>(0.8) | 2<br>(5.7) | 0<br>(0.0) | 2<br>(2.4) | * | * | 2<br>(2.7) | 3<br>(1.6) | 0<br>(0.0) | 0<br>(0.0) | 0<br>(0.0) | 0<br>(0.0) | 0<br>(0.0) | 0<br>(0.0) | 0<br>(0.0) | 9<br>(1.4) |
| Network Size, Mean (SD) | 6.8<br>(4.2) | 6.8<br>(3.9) | 6.1<br>(3.6) | 5.8<br>(3.7) | 6.9<br>(4.14) | 7.0<br>(4.5) | 8.2<br>(4.6) | 4.5<br>(0.7) | 7.4<br>(4.3) | 7.2<br>(4.0) | 7.1<br>(3.9) | 5.6<br>(3.5) | 5.4<br>(3.4) | 7.2<br>(3.2) | 7.4<br>(4.8) | 7.3<br>(5.1) | 6.6<br>(4.3) | 6.7<br>(3.7) |
| PDDS, Mean (SD) | 1.9<br>(2.1) | N/A | 2.2<br>(2.2) | N/A | 2.1<br>(1.9) | N/A | 1.6<br>(2.0) | N/A | 1.8<br>(2.3) | N/A | 1.4<br>(1.9) | N/A | 2.2<br>(2.4) | N/A | 1.6<br>(2.0) | N/A | 1.3<br>(1.9) | N/A |
| MSRS-R, Mean (SD) | 7.6<br>(5.5) | N/A | 8.0<br>(5.5) | N/A | 8.8<br>(5.5) | N/A | * | N/A | 7.4<br>(5.9) | N/A | 6.7<br>(5.6) | N/A | 8.7<br>(5.6) | N/A | 7.0<br>(5.0) | N/A | 6.5<br>(5.8) | N/A |
| PROMIS Physical<br>Function T-score, Mean<br>(SD) | 46.4<br>(10.8) | 56.3<br>(9.0) | 45.3<br>(10.6) | 51.2<br>(10.4) | 45.8<br>(11.5) | 53.5<br>(8.9) | 48.9<br>(10.3) | 42.7<br>(4.5) | 46.1<br>(11.9) | 59.2<br>(8.0) | 47.3<br>(10.3) | 45.5<br>(10.9) | 43.7<br>(10.6) | 45.5<br>(7.8) | 47.7<br>(10.8) | 42.3<br>(11.2) | 48.3<br>(9.2) | 57.5<br>(7.9) |

- a. Cross-sectional cohorts include Cohort 1 and 2 (University of Pittsburgh), Cohort 3 and 4 (Columbia University), Cohort 5 (Yale University), Cohort 6 (University of Buffalo Medical Center), Cohort 7 (University of Pennsylvania), and Cohort 8 (GEMS cohort) - b. PDDS: Patient-Determined Disease Steps. - c. MSRS-R: Multiple Sclerosis Rating Scale-Revised. - d. PROMIS: Patient-Reported Outcomes Measurement Information System Physical Function. - e. Abbreviation: \* - Data unavailable, N/A - not applicable.

**eTable 2.** Cross-sectional analysis of personal social network features in relation to PROMIS Physical Function during the COVID-19 pandemic in control participants.

|  | N | Beta <sup>a</sup> | 95% CI<br>(lower) | 95% CI<br>(upper) | p value <sup>b</sup> |
| --- | --- | --- | --- | --- | --- |
| *Network Structure* |  |  |  |  |  |
| Size | 1047 | 0.172 | 0.048 | 0.295 | .007 |
| Density | 1005 | -0.693 | -2.689 | 1.304 | .496 |
| Constraint | 1005 | -0.034 | -0.063 | -0.005 | .020 |
| Effective Size | 1005 | 0.313 | 0.018 | 0.608 | .038 |
| Maximum Degree | 1005 | 0.383 | 0.141 | 0.626 | **.002** |
| Mean Degree | 1005 | 0.326 | 0.025 | 0.628 | .034 |
| *Network Composition* |  |  |  |  |  |
| Percent Kin | 1022 | -1.065 | -2.800 | 0.669 | .228 |
| Standard Deviation of Age | 760 | -0.025 | -0.123 | 0.073 | .619 |
| Diversity of Sex | 1022 | 0.596 | -1.021 | 2.212 | .470 |
| Diversity of Race | 1020 | 2.844 | -0.013 | 5.702 | .051 |
| Percent contacted weekly or less | 1022 | -1.522 | -3.840 | 0.796 | .198 |
| Percent known for less than 6 years | 1022 | 0.713 | -1.581 | 3.007 | .542 |
| Percent who live over 15 miles away | 1022 | -0.741 | -2.541 | 1.059 | .419 |
| Percent who drink | 1022 | -1.138 | -2.917 | 0.642 | .210 |
| Percent who smoke | 1022 | -4.846 | -7.919 | -1.774 | **.002** |
| Percent non exercisers | 1022 | -1.574 | -3.212 | 0.064 | .060 |
| Percent bad diet | 1022 | -2.556 | -4.267 | -0.845 | .003 |
| Percent who have a negative health influence | 1022 | -5.707 | -7.405 | -4.010 | **<.001** |

- a. Adjusted for potential confounders including age, employment, and income. Disease duration was not adjusted for since it does not apply to control participants. - b. Bolded p values meet the significance threshold of $p < .002$ ( $\alpha = 0.05$ , corrected for 18 comparisons).

**eTable 3.** Longitudinal examination of the association between changes in personal social network features (pandemic values minus pre-pandemic baseline) in relation to patient-reported outcomes in people with multiple sclerosis during the pandemic.

|  | PDDS <sup>a</sup> |  |  |  |  | MSRS-R <sup>b</sup> |  |  |  |  | PROMIS <sup>c</sup> |  |  |  |  |
| --- | --- | --- | --- | --- | --- | --- | --- | --- | --- | --- | --- | --- | --- | --- | --- |
|  | N | Beta <sup>d</sup> | 95% CI (lower) | 95% CI (upper) | p value <sup>d</sup> | N | Beta <sup>d</sup> | 95% CI (lower) | 95% CI (upper) | p value <sup>d</sup> | N | Beta <sup>d</sup> | 95% CI (lower) | 95% CI (upper) | p value <sup>d</sup> |
| ***Network Structure*** |  |  |  |  |  |  |  |  |  |  |  |  |  |  |  |
| Size | 185 | -0.004 | -0.063 | 0.055 | .892 | 185 | -0.071 | -0.211 | 0.070 | .324 | 178 | 0.040 | -0.253 | 0.334 | .786 |
| Density | 166 | -0.571 | -1.951 | 0.809 | .415 | 166 | -1.715 | -5.074 | 1.645 | .315 | 162 | 5.581 | -0.851 | 12.014 | .088 |
| Constraint | 166 | -0.009 | -0.026 | 0.008 | .300 | 166 | -0.012 | -0.053 | 0.029 | .574 | 162 | 0.084 | 0.004 | 0.165 | .041 |
| Effective Size | 166 | 0.027 | -0.172 | 0.225 | .791 | 166 | 0.020 | -0.464 | 0.504 | .935 | 162 | -0.610 | -1.554 | 0.333 | .203 |
| Maximum Degree | 166 | 0.011 | -0.122 | 0.143 | .874 | 166 | 0.005 | -0.317 | 0.327 | .975 | 162 | -0.248 | -0.88 | 0.385 | .440 |
| Mean Degree | 166 | 0.012 | -0.156 | 0.179 | .891 | 166 | -0.055 | -0.464 | 0.354 | .791 | 162 | -0.234 | -1.048 | 0.58 | .571 |
| ***Network Composition*** |  |  |  |  |  |  |  |  |  |  |  |  |  |  |  |
| Percent Kin | 171 | -0.952 | -2.225 | 0.320 | .141 | 171 | -0.849 | -3.956 | 2.257 | .590 | 166 | 7.697 | 1.737 | 13.657 | .012 |
| Standard Deviation of Age | 139 | -0.069 | -0.159 | 0.022 | .129 | 139 | -0.137 | -0.399 | 0.125 | .290 | 134 | 0.016 | -0.496 | 0.528 | .945 |
| Diversity of Sex | 170 | 0.181 | -0.728 | 1.089 | .695 | 170 | -1.540 | -3.700 | 0.621 | .161 | 165 | -1.049 | -5.361 | 3.262 | .631 |
| Diversity of Race | 170 | -1.456 | -3.754 | 0.842 | .213 | 170 | -3.522 | -9.164 | 2.119 | .219 | 165 | 3.909 | -7.267 | 15.085 | .490 |
| Percent contacted weekly or less | 171 | 0.183 | -1.103 | 1.469 | .779 | 171 | -0.442 | -3.562 | 2.679 | .780 | 166 | -2.732 | -8.807 | 3.343 | .375 |
| Percent known for less than 6 years | 171 | 1.080 | -0.569 | 2.729 | .198 | 171 | 1.071 | -2.949 | 5.091 | .599 | 166 | -5.856 | -13.668 | 1.956 | .141 |
| Percent who live over 15 miles away | 171 | -0.396 | -1.319 | 0.527 | .398 | 171 | -0.901 | -3.141 | 1.339 | .428 | 166 | 2.366 | -2.188 | 6.921 | .306 |
| Percent who drink | 143 | -0.56 | -1.938 | 0.818 | .423 | 143 | -1.89 | -4.973 | 1.192 | .227 | 143 | 2.833 | -3.713 | 9.379 | .393 |
| Percent who smoke | 171 | 0.475 | -0.904 | 1.854 | .497 | 171 | -3.319 | -6.626 | -0.012 | .049 | 166 | 0.514 | -6.179 | 7.207 | .880 |
| Percent non exercisers | 171 | 0.439 | -0.394 | 1.271 | .299 | 171 | 1.025 | -0.995 | 3.045 | .318 | 166 | 2.13 | -1.831 | 6.09 | .290 |
| Percent bad diet | 143 | 0.202 | -1.072 | 1.476 | .754 | 143 | -0.434 | -3.294 | 2.427 | .764 | 143 | 2.418 | -3.626 | 8.461 | .430 |
| Percent who have a negative health influence | 143 | 0.113 | -0.852 | 1.077 | .817 | 143 | 0.253 | -1.912 | 2.418 | .817 | 143 | -1.144 | -5.725 | 3.437 | .622 |

- a. PDDS: Patient-Determined Disease Steps. - b. MSRS-R: Multiple Sclerosis Rating Scale-Revised. - c. PROMIS: Patient-Reported Outcomes Measurement Information System Physical Function. - d. Adjusted for potential confounders including age, disease duration, employment, income, study cohort, and time lapse between pre-pandemic and pandemic assessment. None of the p-values meet the significance threshold of $p < .00092$ ( $\alpha = 0.05$ , corrected for 54 comparisons).

**eTable 4.** Moderation analysis assessing the influence of multiple sclerosis diagnosis on the direction and strength of the association between personal social network features and PROMIS Physical Function during the COVID-19 pandemic.

|  |  | Association between Network Feature and PROMIS Physical Function |  |  |  | Moderating Effect of MS Diagnosis on Association between Network Feature and PROMIS Physical Function |  |  |  |
| --- | --- | --- | --- | --- | --- | --- | --- | --- | --- |
|  | N | Beta | 95% CI (lower) | 95% CI (upper) | p value <sup>a</sup> | Beta | 95% CI (lower) | 95% CI (upper) | p value <sup>a</sup> |
| *Network Structure* |  |  |  |  |  |  |  |  |  |
| Size | 1985 | 0.194 | -0.112 | 0.5 | .214 | 0.035 | -0.157 | 0.226 | .723 |
| Density | 1896 | -1.047 | -6.032 | 3.938 | .680 | 0.389 | -2.77 | 3.547 | .809 |
| Constraint | 1896 | -0.067 | -0.138 | 0.004 | .064 | 0.022 | -0.022 | 0.067 | .325 |
| Effective Size | 1896 | 0.423 | -0.325 | 1.17 | .267 | -0.018 | -0.502 | 0.466 | .943 |
| Maximum Degree | 1896 | 0.784 | 0.179 | 1.389 | .011 | -0.27 | -0.654 | 0.115 | .169 |
| Mean Degree | 1896 | 0.709 | -0.036 | 1.454 | .062 | -0.282 | -0.745 | 0.182 | .233 |
| *Network Composition* |  |  |  |  |  |  |  |  |  |
| Percent Kin | 1927 | -0.234 | -4.554 | 4.085 | .915 | -0.924 | -3.656 | 1.808 | .507 |
| Standard Deviation of Age | 1321 | -0.019 | -0.258 | 0.22 | .876 | 0.002 | -0.147 | 0.151 | .976 |
| Diversity of Sex | 1921 | 0.915 | -3.117 | 4.947 | .656 | -0.351 | -2.925 | 2.22 | .789 |
| Diversity of Race | 1906 | 8.788 | 1.409 | 16.167 | .019 | -5.878 | -10.889 | -0.868 | **.022** |
| Percent contacted weekly or less | 1927 | -0.547 | -6.354 | 5.259 | .853 | 0.329 | -3.407 | 4.064 | .863 |
| Percent known for less than 6 years | 1927 | -3.821 | -9.437 | 1.796 | .182 | 2.744 | -0.989 | 6.477 | .150 |
| Percent who live over 15 miles away | 1927 | -5.434 | -9.91 | -0.957 | .017 | 4.662 | 1.811 | 7.512 | **.001** |
| Percent who drink | 1927 | -5.679 | -10.262 | -1.096 | .015 | 4.697 | 1.653 | 7.741 | **.002** |
| Percent who smoke | 1927 | -7.724 | -15.198 | -0.25 | .043 | 1.136 | -3.451 | 5.723 | .627 |
| Percent non exercisers | 1927 | -3.508 | -7.576 | 0.56 | .091 | 1.116 | -1.449 | 3.682 | .394 |
| Percent bad diet | 1927 | -1.633 | -5.924 | 2.658 | .456 | -0.972 | -3.766 | 1.821 | .495 |
| Percent who have a negative health influence | 1927 | -11.028 | -15.237 | -6.819 | <.001 | 4.427 | 1.786 | 7.069 | **.001** |

a. Bolded p-values met the significance threshold of $p < .05$ .

**eTable 5.** Moderation analysis assessing the influence of multiple sclerosis diagnosis on the direction and strength of the association between personal social network features and PROMIS Physical Function during the COVID-19 pandemic in people with multiple sclerosis and control participant subgroups.

|  | MS |  | Control |  |
| --- | --- | --- | --- | --- |
|  | Slope [95% CI] | p-value | Slope [95% CI] | p-value |
| Diversity of Race | -2.968 [-6.893, 0.956] | .138 | 2.910 [-0.224, 6.044] | .068 |
| Percent who live over 15 miles away | 3.890 [1.852, 5.928] | **<.001** | -0.772 [-2.765, 1.221] | .448 |
| Percent who drink | 3.716 [1.392, 6.039] | **.002** | -0.982 [-2.966, 1.003] | .332 |
| Percent who have a negative health influence | -2.174 [-4.019, -0.329] | **.021** | -6.601 [-8.494, -4.708] | **<.001** |

a. Bolded p-values met the significance threshold of $p < .05$ .

